# Schwann cells contribute to keloid formation

**DOI:** 10.1101/2021.08.09.21261701

**Authors:** Martin Direder, Tamara Weiss, Dragan Copic, Vera Vorstandlechner, Maria Laggner, Caterina Selina Mildner, Katharina Klas, Daniel Bormann, Werner Haslik, Christine Radtke, Matthias Farlik, Lisa Shaw, Bahar Golabi, Erwin Tschachler, Konrad Hötzenecker, Hendrik Jan Ankersmit, Michael Mildner

**Affiliations:** Laboratory for Cardiac and Thoracic Diagnosis, Regeneration and Applied Immunology, Department of Thoracic Surgery, Medical University of Vienna, Vienna, Austria; Aposcience AG (FN 308089y), Dresdner Straße 87/A21, Vienna, Austria; Department of Plastic, Reconstructive and Aesthetic Surgery, Medical University of Vienna, Vienna, Austria; Department of Obstetrics and Gynecology, Medical University of Vienna, Vienna, Austria; Department of Dermatology, Medical University of Vienna, Vienna, Austria; Department of Thoracic Surgery, Medical University of Vienna, Vienna, Austria

## Abstract

Keloids are disfiguring, hypertrophic scars with yet poorly understood pathomechanisms, which could lead to severe functional impairments. Here we analyzed the characteristics of keloidal cells by single cell sequencing and discovered the presence of an abundant population of Schwann cells that persisted in the hypertrophic scar tissue after wound healing. In contrast to normal skin, keloidal Schwann cells possess a repair-like phenotype and high cellular plasticity. Our data support the hypothesis that keloidal Schwann cells contribute to the formation of the extracellular matrix and are able to affect M2 polarization of macrophages. Indeed, we show that macrophages in keloids predominantly display a M2 polarization and produce factors that inhibit Schwann cell differentiation. Our data suggest a contribution of this cross-talk to the continuous expansion of keloids, and that targeting Schwann cells might represent an interesting novel treatment option for keloids.

## Introduction

Keloids are fibroproliferative, protruding scar-like pathologies of the skin^1^ characterized by a persisting, gradual growth beyond the margin of the wound into the surrounding healthy skin. In susceptible individuals even minor skin injuries, such as insect bites or vaccinations, can induce keloid formation^2,3^. Although keloids show some tumor-like behavior, they do not metastasize. Nonetheless, keloids can cause severe pain, chronic pruritus, psychosocial impairment and movement restriction due to their scar-like character^3-6^. Genetic predispositions as well as chronic inflammatory processes are being discussed in disease etiology^2,6-8^. As keloids are characterized by an increased proliferation of fibroblasts and extensive over-production of ECM components, keloid research has primarily focused on the involvement of fibroblasts in the development of these lesions^1^. Although these studies have identified numerous potentially pathology-related factors, the fundamental patho-mechanistic events driving keloid formation remain unclear^1^. The limited treatment options include steroid injections, *γ*-radiation, and surgery, but the majority of patients still suffer from high recurrence rates^4,9,10^, which underline the importance of identifying novel therapeutic approaches.

Due to the pruritic and painful nature of keloids, a neuronal contribution to the pathogenesis is conceivable. The subepidermal nerve plexus of the skin is the largest sensory organ of the human body. There is growing evidence that cutaneous innervation plays an important role in mediating wound healing^15,16^. It is, thus, surprising that a contribution of cutaneous nerves in keloid formation is poorly investigated^11-14^. The major cellular constituents of peripheral nerves are Schwann cells. In the healthy skin, Schwann cells ensheath cutaneous axons and ensure the integrity and function of sensory neurons^17-19^. Recently, increasing attention was drawn to Schwann cells because of their ability to adopt a transient repair phenotype in response to peripheral nerve injury^20,21^. This process involves a de-differentiation step into a proliferative precursor or immature-like Schwann cell state and the acquisition of repair specific functions ^22^. These dedicated repair Schwann cells express (neuro)trophic factors to support neuronal survival and form regeneration tracks (Bungner bands) to promote axonal outgrowth and guidance^23,24^. Furthermore, repair Schwann cells release a plethora of chemokines and cytokines to attract macrophages, thereby contributing to clear the lesion from myelin debris and remodelling the ECM to facilitate nerve regeneration^25-28^. Previous studies further support that the interaction between Schwann cells and macrophages affect their phenotype. While macrophages are known to regulate repair Schwann cell re-differentiation, Schwann cells promote the induction of M2 polarization of macrophages^29^. Macrophages play a crucial role in cutaneous wound healing by modulating the microenvironment during the different healing stages^30-32^. Especially M2-macrophages are associated with fibrosis and scarring and persist in keloids^31,33,34^. Moreover, a recent study in mice reported that the wound microenvironment is a key determinant of Schwann cell behavior, influencing their proliferation status, re-programming into mesenchymal-like cells, immune signaling and ECM production^35^. Indeed, Schwann cells have been shown to contribute not only to nerve regeneration but also to wound healing by regulating myofibroblast differentiation, epithelial proliferation and ECM formation^36^. Hence, Schwann cells are important players in cutaneous wound healing processes and might play an as yet underappreciated role in fibrotic processes.

In the present study we performed single-cell RNA sequencing (scRNAseq) of keloids to analyze the entire cellular spectrum and the transcriptional landscape at a single cell resolution and to identify hitherto underestimated cellular and molecular players of keloid pathogenesis. Our analysis revealed a yet not described population of highly plastic keloidal Schwann cells. The vast majority of keloidal Schwann cells was not associated with axons, displayed a de-differentiated, repair-like phenotype, and showed key features with a high potential for affecting ECM deposition and macrophage function. Our findings suggest that an abnormal reaction of Schwann cells to skin injuries contribute to keloid pathogenesis.

## Results

### scRNAseq reveals enrichment of Schwann cell in keloids

To investigate the cellular composition of keloids, we performed scRNAseq and compared our data with a recently published scRNAseq data set of normal human skin^37^. In total, transcriptomic data of 19598 cells from normal skin and 47478 cells from keloids were analyzed. After unbiased cluster generation (Fig. 1a), cell clusters were identified with well-established marker genes as well as computed clustermarkers (Supplementary Figs. 1a and 1b; Supplementary Table 1). Cell clusters have been assigned to fibroblasts (FB), smooth muscle cells and pericytes (SMC/PC), keratinocytes (KC), endothelial cells (EC), lymphatic endothelial cells (LEC), T-cells (TC), macrophages (MAC), dendritic cells (DC), melanocytes (MEL) erythrocytes (ERY) and Schwann cells (SC), (Fig. 1a). Comparison of different cell types between normal skin and keloids revealed a significant increase in cell number of fibroblasts, endothelial cells and lymphatic endothelial cells^1,38^ (Fig.1a). Immunofluorescence (IF) staining corroborated our scRNAseq data by showing an increase in fibrotic (vimentin positive) and well-vascularized (smooth-muscle-actin positive) tissue of keloids compared to healthy skin (Supplementary Fig. 2). Strikingly, relative numbers of Schwann cells (S100B^+^/NGFR^+^) were strongly increased in keloids (Fig. 1b). To compare the SC populations between skin and keloid samples, IF staining for the Schwann cell marker S100B was performed. In healthy skin S100B stained epidermal Langerhans cells, melanocytes and dermal Schwann cells found in the subepidermal nerve plexus, nerve structures around sweat glands, and larger nerve bundles of the deep dermis (Fig. 1c). By contrast, S100B staining of keloids showed a scattered and disorganized distribution of Schwann cells in the dermis (Fig. 1c). Morphologically, Schwann cells in keloids showed an elongated, bipolar shape with a spindle-shaped body (Fig. 1d). To further determine whether Schwann cells were associated with neurons, we stained 100 µm thick transversal dermal sections of normal skin and keloids with S100B and the axon marker PGP9.5. Throughout all dermal layers of healthy skin, Schwann cells were found to ensheath axons (Fig.2a), while the vast majority of Schwann cells in keloids was not associated with axons up to 6 mm depth (Fig. 2b) (Supplementary movies 1-5). Only in the deep dermis of keloids (6000-8500 µm), neuron-ensheathing Schwann cells were detected.

**Figure 1.**
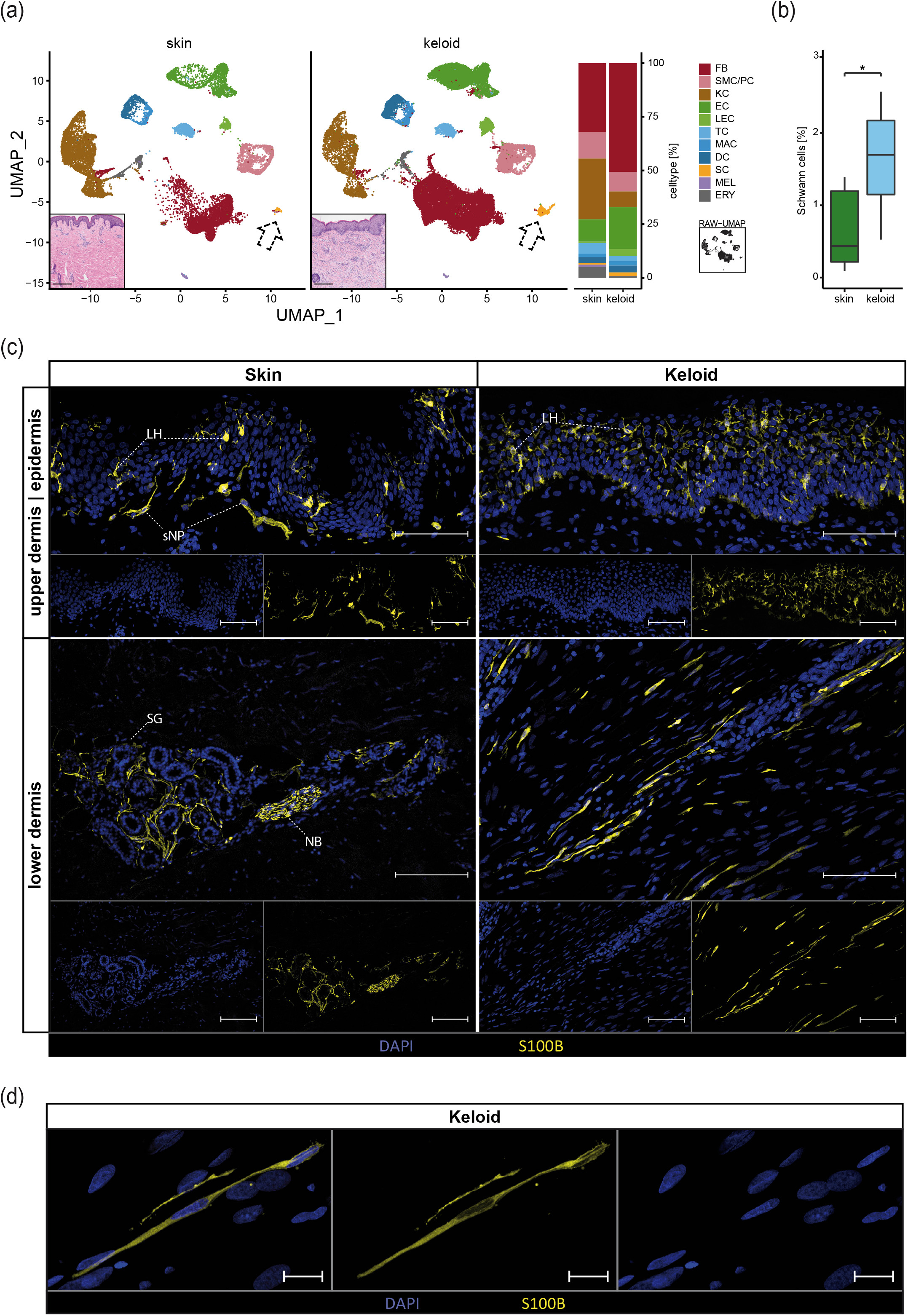
Single cell RNA sequencing revealed Schwann cell accumulation in keloids compared to healthy skin. (a) Integrated UMAP-plots comparing cells of healthy skin (n=7) with keloid tissue (n=4), split by tissue, identifying fibroblasts (FB), smooth muscle cells and pericytes (SMC/PC), keratinocytes (KC), endothelial cells (EC), lymphatic endothelial cells (LEC), T-cells (TC), macrophages (MAC), dendritic cells (DC), Schwann cells (SC), melanocytes (MEL) and erythrocytes (ERY). Bar plot indicates relative amounts of cell types within skin and keloid samples. RAW-UMAP represents unsplit UMAP without cluster colouring. Inserts show representative micrographs of Hematoxylin-Eosin-stained skin and keloid sections. Scale bars: 250 µm; dashed arrowheads indicate SC-cell clusters. (b) Comparison of relative SC amounts by tissue. * indicates p-value<0.05. (c) Representative immunostainings of skin (left-side) and keloid (right-side) with S100B-positive SCs in the dermal area. LH = Langerhans cell, sNP = subepidermal nerve plexus, SG = sweat gland, NB = nerve bundle. Scale bars: 100 µm; (d) Close-up of keloidal SCs; Scale bar: 20 µm. Tissues of n = 3 donors were analysed per staining.

**Figure 2.**
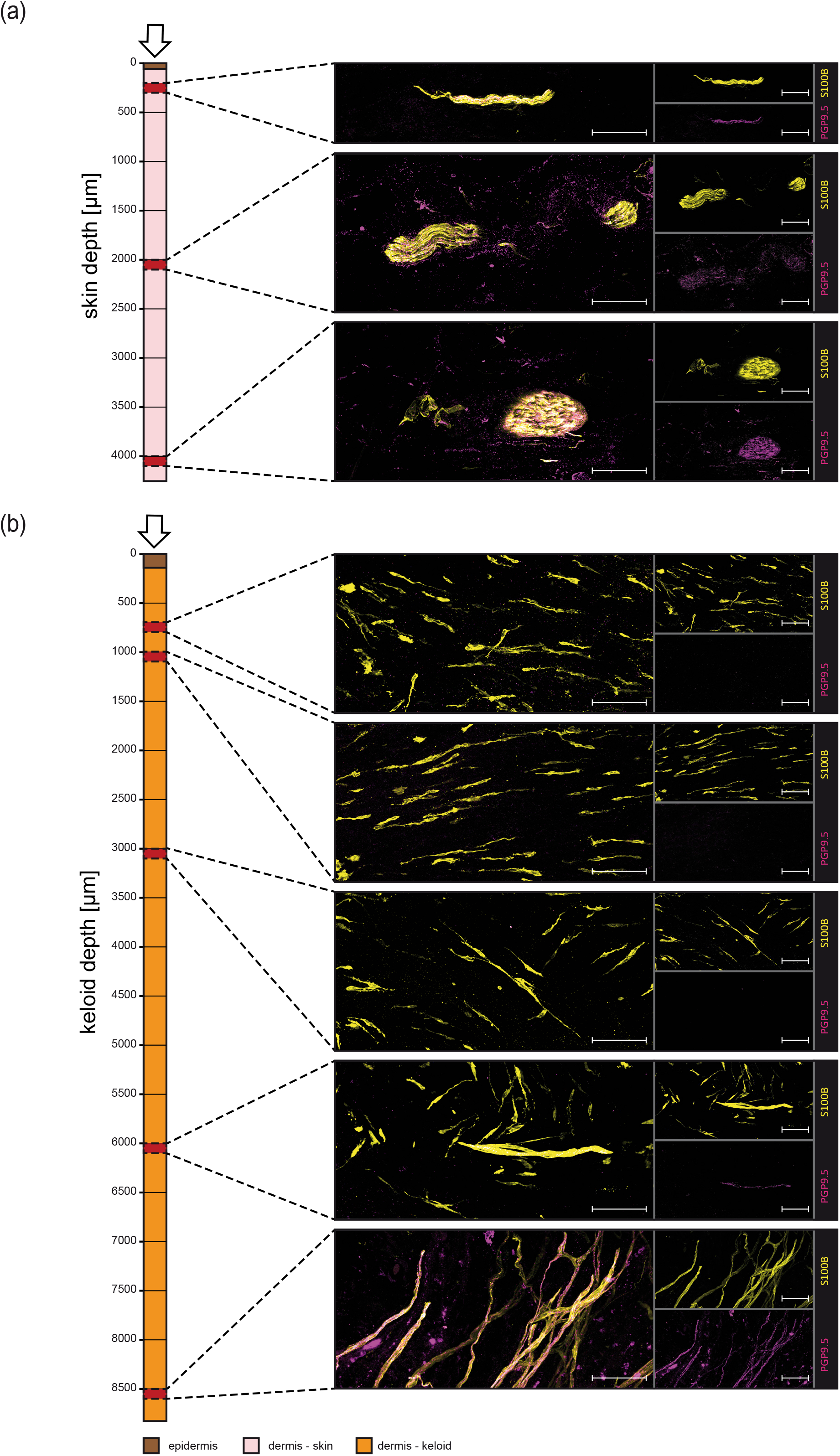
Nerve-autonomous Schwann cell distribution throughout the whole keloidal dermis. Bird’s-eye view of 100 µm-thick sections of (a) healthy skin and (b) keloids dermal sheets. S100B indicates Schwann cells and PGP 9.5 nerve fibers. Vertical reference bar illustrates depth of the depicted sheet region below the skin surface. Scale bar: 100 µm.

### Schwann cells present in keloids show high cellular plasticity

To characterize the Schwann cell population in keloids in more detail, we performed subclustering and detected four Schwann cell subtypes exclusively present in keloids (SC-Repair, SC-EC, SC-FB and SC-Prolif). One major Schwann cell subtype, here referred to as SC-Skin was detected specifically in normal skin, and one Schwann cell subtype referred to as SC-Promyel was present in both normal skin and keloids (SC-Promyel) (Fig. 3a and Supplementary Fig. 3a). A total of 370 genes were differentially expressed between skin- and all keloid-derived Schwann cells (144 up- and 226 down-regulated) (Fig. 3b). The skin-specific Schwann cell cluster (SC-Skin) expressed several genes characteristic for myelinating Schwann cells [myelin basic protein (*MBP*), proteolipid protein (*PLP1*), peripheral myelin protein 22 (*PMP22*) and myelin protein zero (*MPZ*)] as well as genes commonly expressed in non-myelinating Schwann cells [neural cell adhesion molecule 1 (NCAM1), L1 cell adhesion molecule (L1CAM)]^23^ (Fig. 3c). The Schwann cell cluster present in both tissues was enriched in the keloids, presumably representing pro-myelinating Schwann cells (SC-Promyel), characterized by reduced expression levels of *MPZ* and *PLP1* and no *MBP* (Fig. 3c). In line with our scRNAseq data, double staining of S100B with MBP confirmed that myelinating Schwann cells were present in healthy skin but absent from keloids (Fig. 5a).

**Figure 3.**
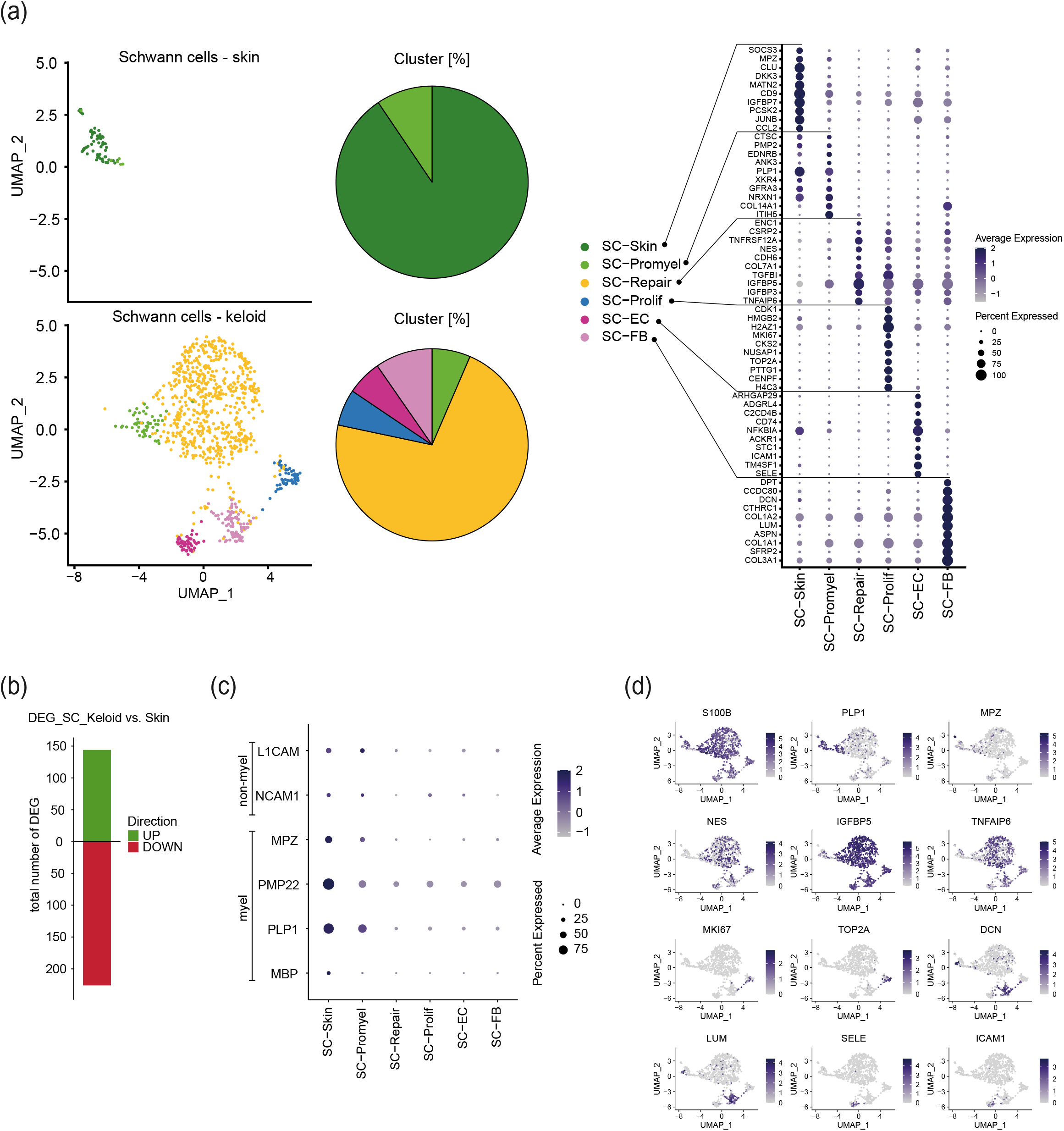
Identification of keloid-specific Schwann cell subtypes. (a) Analysis of the Schwann cell subset identified 6 distinct Schwann cell subtypes depicted in UMAP-plots split by tissue. Pieplots display cluster percentage of the identified cell subtypes within each condition. Dotplot shows the top 10 genes leading to subcluster formation. Identified Schwann cell cluster: Myelinating and non-myelinating Schwann cells of the healthy skin (SC-Skin), promyelinating Schwann cells (SC-Promyel), repair Schwann cells (SC-Repair), proliferating Schwann cells (SC-Prolif), cells expressing Schwann cell and endothelial cell specific genes (SC-EC), cells expressing Schwann cell and fibroblast specific genes (SC-FB). Colour codes indicate average gene expression levels; Dot sizes indicate relative amounts of positive cells. (b) Total amount of differentially expressed genes (DEG) of all keloid-Schwann cells compared to all skin-Schwann cells. Genes with an average log-foldchange ≥ 2 were included. (c) Mature Schwann cell assignment by validated marker genes; myel = myelinating Schwann cell marker; non-myel = marker genes for non-myelinating Schwann cells. Colour intensity indicates average gene expression. Dot-size shows percentage of cells expressing the genes. (d) Feature Plots of the integrated Schwann cells from healthy skin and keloids shows the expression of S100 calcium binding protein B (*S100B*), proteolipid protein 1 (*PLP1*), myelin protein zero (*MPZ*), nestin (*NES*), insulin-like growth factor binding protein 5 (*IGFBP5*), tumor necrosis factor alpha induced protein 6 (*TNFAIP6*), marker of proliferation Ki-67 (*MKI67*), DNA topoisomerase II alpha (*TOP2A*), decorin (*DCN*), lumican (*LUM*), E-selectin (*SELE*) and intercellular adhesion molecule 1 (*ICAM1*). Normal log gene expressions are mapped on the UMAP-Plot.

The majority of Schwann cells present in keloids highly expressed nestin (*NES*), insulin-like growth factor-binding protein 3 (*IGFBP3*), insulin-like growth factor-binding protein 5 (*IGFBP5*), transforming growth factor beta–induced (*TGFBI*), TNF-alpha induced protein 6 (*TNFAIP6)* and cellular communication network factor 3 (*CCN3*) (Fig. 3a and 3d) ^21,39-41^. To validate our transcriptomics data on the protein level, we performed double staining of S100B and nestin, visualizing that keloidal Schwann cells were highly positive for nestin (Fig. 4a). Nestin is a known marker for neural precursor cells, involved in neuronal/glial development, which is also upregulated in human repair Schwann cells ^42,43^. To characterize the cellular state of keloidal Schwann cells in more detail, we conducted IF staining for SRY-box transcription factor 10 (SOX10) and nerve growth factor receptor (NGFR), which are both known to be upregulated in immature/de-differentiated Schwann cells, as well as the transcription factor JUN which was demonstrated to be a key factor determining the repair identity of Schwann cells ^21^. NGFR was strongly expressed by keloidal Schwann cells but also weakly expressed by vascular cells (Fig. 4b). SOX10 nuclear expression was exclusively found in keloidal Schwann cells (Fig.4c). Of note, keloidal Schwann cell nuclei were positive for c-JUN (Fig. 4d) but c-JUN expression was also found in other cells. Based on the characteristic elongated keloidal Schwann cell morphology together with the expression of markers associated with de-differentiation and repair, the major Schwann cell type in the keloids could be assigned to a repair-like cellular state (SC-Repair).

**Figure 4.**
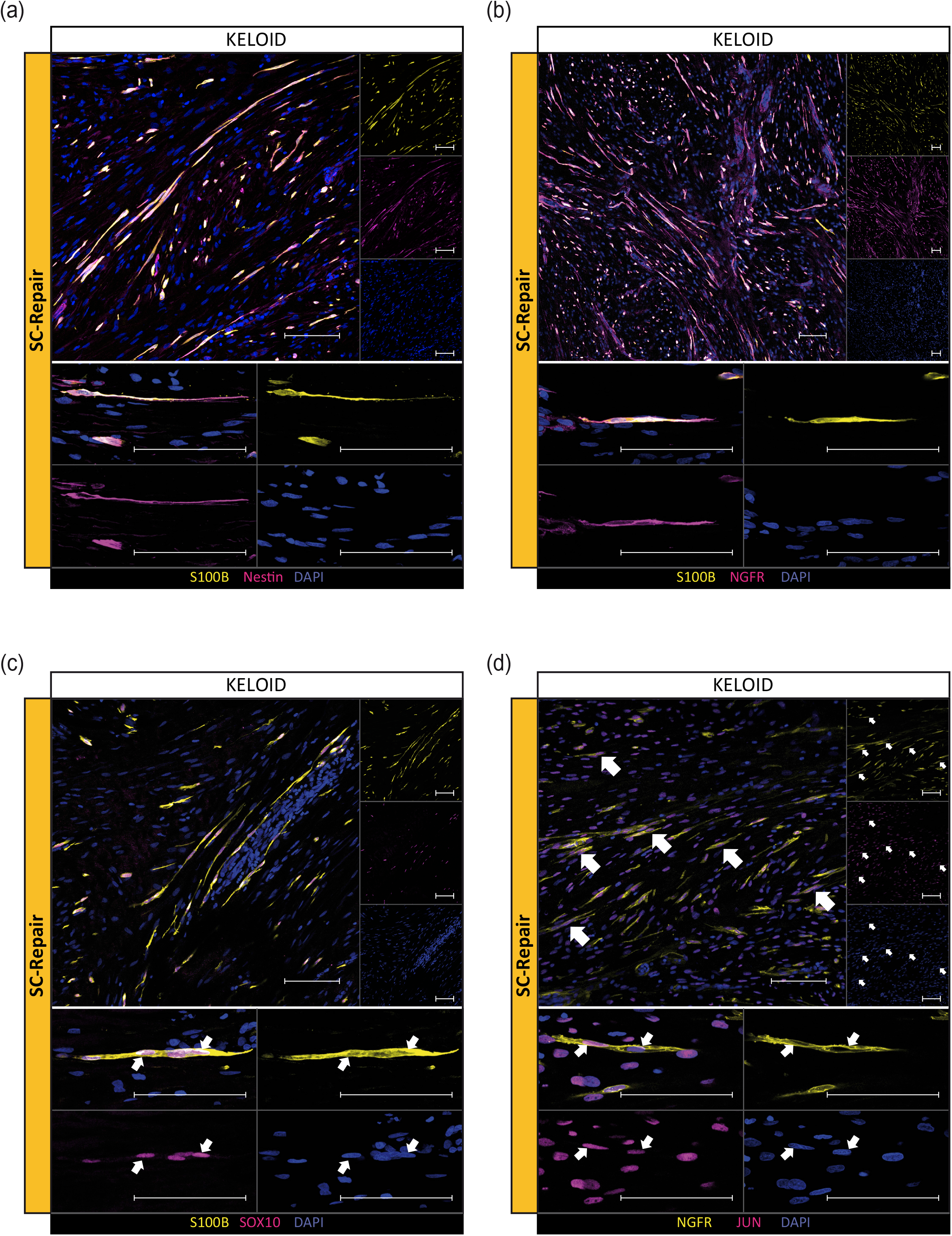
Immunofluorescence staining confirmed predicted Schwann cell subtypes in keloid tissue. Immunostainings of keloidal Schwann cells for (a) S100B and Nestin, (b) S100B and nerve growth factor receptor (NGFR), (c) S100B and SRY-box transcription factor 10 (SOX10) and (d) NGFR and Jun proto-oncogene (JUN). Scale bar: 100 µm. One representative micrograph of n = 3 donors per condition is shown.

In addition, we detected a SC cluster (SC-Prolif) with high expression of genes associated with cell division, such as marker of proliferation Ki-67 (*MKI67*) and DNA-Topoisomerase 2-alpha (*TOP2A)* exclusively in keloids but not in normal skin (Fig. 3a and 3d). IF staining of keloids with KI-67 in combination with NGFR, confirmed the presence of proliferating Schwann cells *in situ*, indicating that keloidal Schwann cells have the ability to re-enter the cell-cycle (Fig. 5c), corresponding to the increased proliferation rates of Schwann cells reported during wound healing and repair processes ^35,36^

**Figure 5.**
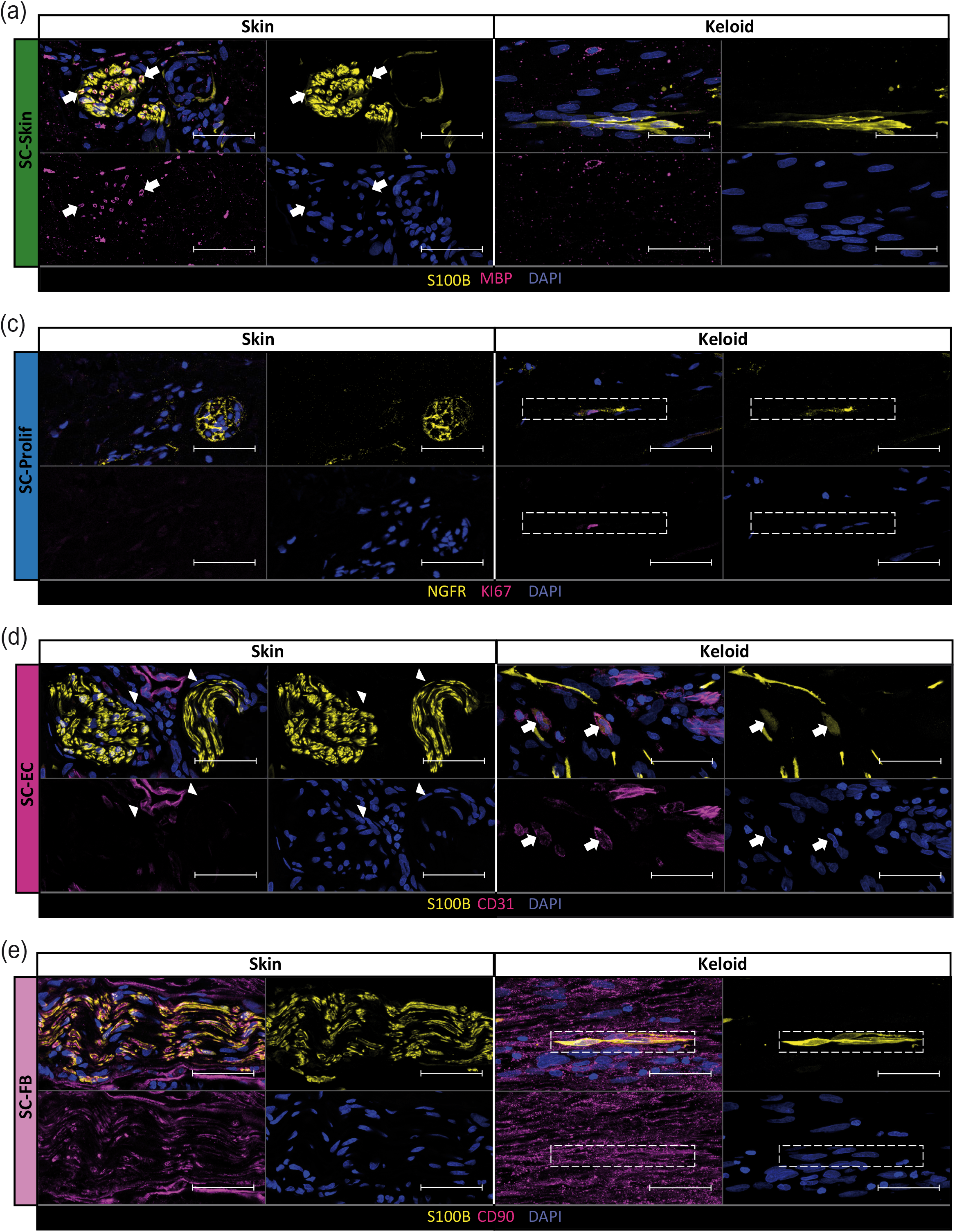
Immunofluorescence staining confirmed predicted Schwann cell subtypes in keloid tissue. Representative immunofluorescence images of Schwann cells double-positive for (a) myelin basic protein (MBP) and S100B in healthy skin and lack of this double positive Schwann cells in the keloid. Representative immunostainings of Schwann cells double positive for (b) nerve growth factor receptor (NGFR) and marker of proliferation Ki-67 (KI67), (d) S100B and cluster of differentiation 31 (CD31) and (e) S100B and THY-1 cell surface antigen (CD90) in the keloid compared to healthy skin. Arrows and rectangles indicate double-positive cells. Arrowheads indicate vessels. Tissues of n = 3 donors per condition were stained. Scale bars: 50 µm.

Two smaller keloid specific Schwann cell populations were characterized in our scRNAseq analysis. One cluster showed a combined expression of Schwann cell and endothelial cell markers [SC-EC; selectin-E (*SELE*) and intercellular adhesion molecule 1 (*ICAM1*)] the other cluster showed a combined expression of Schwann cell and fibroblast markers [SC-FB; lumican (*LUM*), decorin (*DCN*) or THY-1 cell surface antigen (*CD90*)] (Figs. 3a, 3d and Supplementary Figs. 4 and 5). IF stainings confirmed our single cell data and showed double positive cells for S100B and CD31 (SC-EC; Fig. 5d) as well as S100B and CD90 in keloids (SC-FB; Fig. 5e). In healthy skin, CD90-positivity was only observed in axons but not in Schwann cells (Fig. 5e)^44^. Compared to all other S100B-positive Schwann cells, SC-EC showed a different morphology, as they were oval shaped without extensions (Fig. 5d). The SC-FB and SC-EC cell populations could represent an intermediate stage between Schwann cells and fibroblasts or endothelial cells, suggesting that Schwann cells in keloids show a high plasticity and might be able to trans-differentiate into other cell types.

### *In silico* analysis of the Schwann cell differentiation behaviour in keloids

Upon peripheral nerve injury, non- and myelinating Schwann cells start to de-differentiate, regain migratory and proliferative properties, and perform specific repair functions to support the regeneration of damaged nerves.^23,36^ To investigate whether similar processes are obvious during keloid development, we performed pseudotime trajectory analysis. Pseudotime trajectory suggested that keloidal Schwann cells originate from differentiated myelinated Schwann cells (Fig. 6a). Moreover, our calculation indicated that some of these repair Schwann cells acquired a proliferative state or de-differentiated into fibroblasts-like or endothelial-like Schwann cells (Fig. 6a). We further investigated changes in Schwann cell gene expression along the pseudotime axis. While expression of genes associated with myelination (neuroblast differentiation-associated protein (*AHNAK)*, caveolin 1 (*CAV1)*, cluster of differentiation 9 (*CD9)*, neuronal membrane glycoprotein M6-B (*GPM6B), MBP, MPZ, PLP1* or *PMP22*) decreased over pseudotime (Fig. 6b), the expression of genes associated with Schwann cell precursor cells, repair Schwann cells and nerve regeneration, such as *CCN3*, neurexin 1 (*NRXN1)*, platelet derived growth factor alpha chain (*PDGFA)*, pleiotrophin (*PTN)*, protein tyrosine phosphatase receptor type Z1 (*PTPRZ1)*, proliferation-inducing protein 33 (*SPARCL1)* and zinc finger e-box binding homeobox 2 (*ZEB2)* increased (Fig. 6c)^45-52^. Furthermore, factors associated with cell migration, including calreticulin (*CALR*), *TGFBI* and tenascin c (*TNC)*^53-55^, increased along the pseudotime trajectory. A more complete set of genes regulated along the pseudotime trajectory is provided in supplementary Fig. 6. These findings support the hypothesis that keloidal Schwann cells originate from differentiated myelinated Schwann cells by decreasing the expression of myelin genes while upregulating genes involved in repair mechanisms and migration.

**Figure 6.**
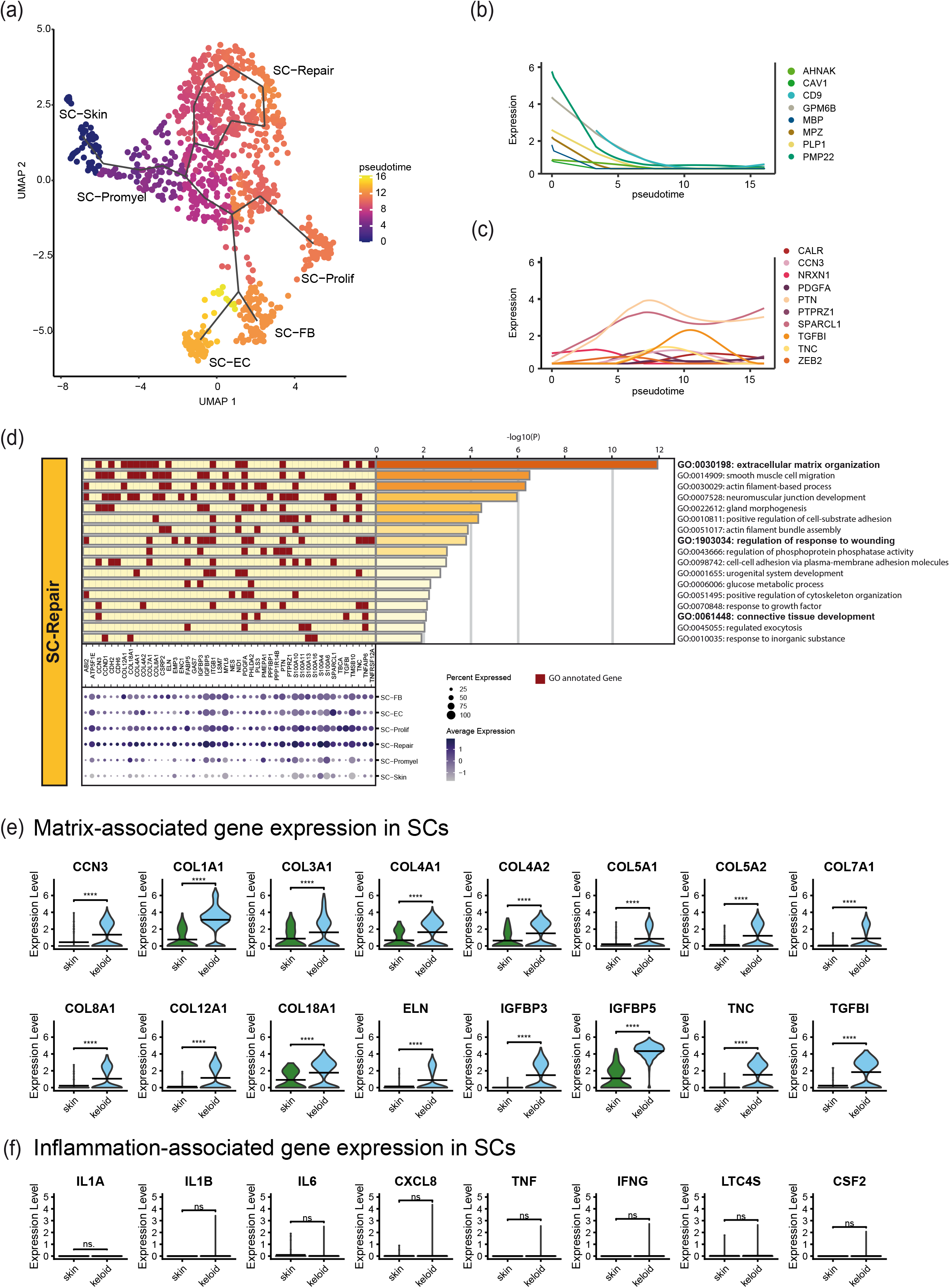
Pseudotime analysis revealed genetic transformation from healthy SCs to keloidal SCs and keloid-specific Repair Schwann cells contribute to ECM formation. (a) UMAP-plot including pseudotime track starting from healthy skin Schwann cells (SC-Skin) to keloidal Schwann cells. Colour code indicates cellular moment in the calculated pseudotime. (b) Regulation of the expression of the myelination-associated genes: myelin basic protein (*MBP*), cluster of differentiation 9 (*CD9*), myelin protein zero (*MPZ*), proteolipid protein 1 (*PLP1*), neuroblast differentiation-associated protein (*AHNAK*), caveolin 1 (*CAV1*), neuronal membrane glycoprotein M6-B (*GPM6B*) and peripheral myelin protein 22 (*PMP22*) along the pseudotime. (c) Expression of genes associated with Schwann cell precursor cells, repair Schwann cells, nerve regeneration and cell migration: zinc finger e-box binding homeobox 2 (*Zeb2*), neurexin 1 (*NRXN1*), proliferation-inducing protein 33 (*SPARCL1*), pleiotrophin (*PTN*), protein tyrosine phosphatase receptor type Z1 (*PTPRZ1*), platelet derived growth factor alpha chain (*PDGFA*), cellular communication network factor 3 (*CCN3*), transforming growth factor beta–induced (*TGFBI*), tenascin C (*TNC*) and calreticulin (*CALR*). Only SC-Skin, SC-Promyel and SC-Repair have been included to the pseudotime-expression analysis. (d) GO-term enrichment of top-clustermarker genes with average log-foldchange ≥ 1.5 from SC-Repair. Bar length represents statistical significance of the annotated term. Genes annotated to the respective term are marked in red. Dotplot depicts expression of the top-clustermarker genes in all Schwann cell subtypes. Colour intensity of the dots depicts average gene expression. Dot size symbolizes percentage of cells expressing the respective genes. (e) Expression of the matrix-associated genes cellular communication network factor 3 (*CCN3*), collagen type I alpha 1 (*COL1A1*), collagen type III alpha 1 (*COL3A1*), collagen type IV alpha 1 (*COL4A1*), collagen type IV alpha 2 (*COL4A2*), collagen type V alpha 1 (*COL5A1*), collagen type V alpha 2 (*COL5A2*), collagen type VII alpha 1 (*COL7A1*), collagen type VIII alpha 1 (*COL8A1*), collagen type XII alpha 1 (*COL12A1*), collagen type XVIII alpha 1 (*COL18A1*), elastin (*ELN*), insulin like growth factor binding protein 3 (*IGFBP3*), insulin like growth factor binding protein 5 (*IGFBP5*), tenascin c (*TNC*) and transforming growth factor beta induced (*TGFBI*) of Schwann cells from the healthy skin and keloid. (f) Expression of the inflammation-associated genes interleukin 1 alpha (*IL1A*), interleukin 1 beta (*IL1B*), interleukin 6 (*IL6*), interleukin 8 (*CXCL8*), tumor necrosis factor alpha (*TNF*), interferon gamma (*IFNG*), leukotriene C4 synthase (*LTC4S*) and colony-stimulating factor 2 (*CSF2*) in Schwann cells of healthy skin and keloids. Crossbeam of violin plots depicts mean expression value. vertical lines show maximum expression. Width represents frequency of cells at the respective expression level; ns. p-value >0.05; ****p-value <0.0001.

### Keloidal Schwann cells contribute to the formation of the extracellular matrix

Pseudotime trajectory analysis indicated functional changes of Schwann cells in keloids. To investigate the transcriptional differences and their possible functional consequences, we utilized GO-Term enrichment analyses. Genes highly expressed in normal skin Schwann cells were strongly associated with myelination and neuron development (Supplementary Fig. 7a), as well as membrane assembly, macrophage chemotaxis and dendritic cell differentiation (Supplementary Fig. 7b). GO-terms of the keloid-specific Schwann cell clusters differed significantly from those of Schwann cells present in normal skin. The most prominent GO-terms found in SC-Prolif were associated with cell proliferation processes (Supplementary Fig. 8a). Genes specifically expressed in SC-EC were mainly associated with the regulation of inflammatory response, and response to bacteria but also with vasculature development and regulation of epithelial cell differentiation (Supplementary Fig. 8b). The gene set enriched in SC-FB showed strong association with processes regulating the production and assembly of the ECM (Supplementary Fig. 8c). Interestingly, genes strongly expressed in SC-Repair were associated with organization of the ECM, response to wound healing and connective tissue development (Fig. 6d). Overall, the expression of many genes associated with ECM production and assembly^56^ was significantly up-regulated in keloids (Supplementary Fig. 9-11) and various matrix-associated genes were enriched specifically in keloidal Schwann cells (Fig. 6e). Since inflammatory processes have been reported to be involved in the pathogenesis of keloids, we further investigated the expression of factors related to skin inflammation. However, the expression of inflammatory mediators was even down-regulated in keloids compared to healthy skin (Supplementary Fig. 12) and not regulated in Schwann cells (Fig. 6f). Together these data indicate a contribution of Schwann cells to the organization of the ECM but not the inflammatory milieu in keloids.

### Schwann cells in keloids differ significantly from Schwann cells found in neurofibroma type 1

Since the majority of Schwann cells in keloids were not associated with axons and displayed a repair-related phenotype, we next explored whether similar Schwann cell populations are found in cutaneous neurofibroma type 1 (NF1), a benign skin-tumor originating from Schwann cells^57^. Therefore, we compared our data set with previously published scRNAseq data of NF1^58^. The re-calculated UMAP (Fig. 7a) and cluster markers (Fig. 7b) differed significantly between SC-NF1, SC-Skin and most keloidal Schwann cells. Only the SC-EC cluster showed high transcriptional resemblance with SC-NF1 (Fig. 7a, left and middle panel; red oval) resulting in a new shared cluster after combined calculation (Fig. 7a, right panel and Fig. 7c). Cells of this combined cluster displayed a transcriptional profile associated with inflammatory processes (Supplementary Fig. 13a). Importantly, no repair-related Schwann cells were found in NF1. In addition, pseudotime trajectory of the combined data set showed that SC-Repair and SC-NF1 represent two separate branches, both originating from myelinating skin Schwann cells (Fig. 7d). Expression of genes associated with matrix formation was decreased in NF1, suggesting that NF1-derived Schwann cells are not involved in tissue remodelling processes (Fig. 7e). However, expression of several inflammatory mediators, such as interleukin 6 (*IL6*), interleukin 8 (*CXCL8*), nuclear receptor 4A1 (*NR4A1*) and nuclear receptor 4A2 (*NR4A2*) was strongly upregulated in the NF1-derived Schwann cells (Fig. 7f), indicating that they contribute to tissue inflammation. Hence, our comparison shows that Schwann cells in keloids and Schwann cells in NF1 differ drastically in their ability to affect tissue remodeling and inflammation.

**Figure 7.**
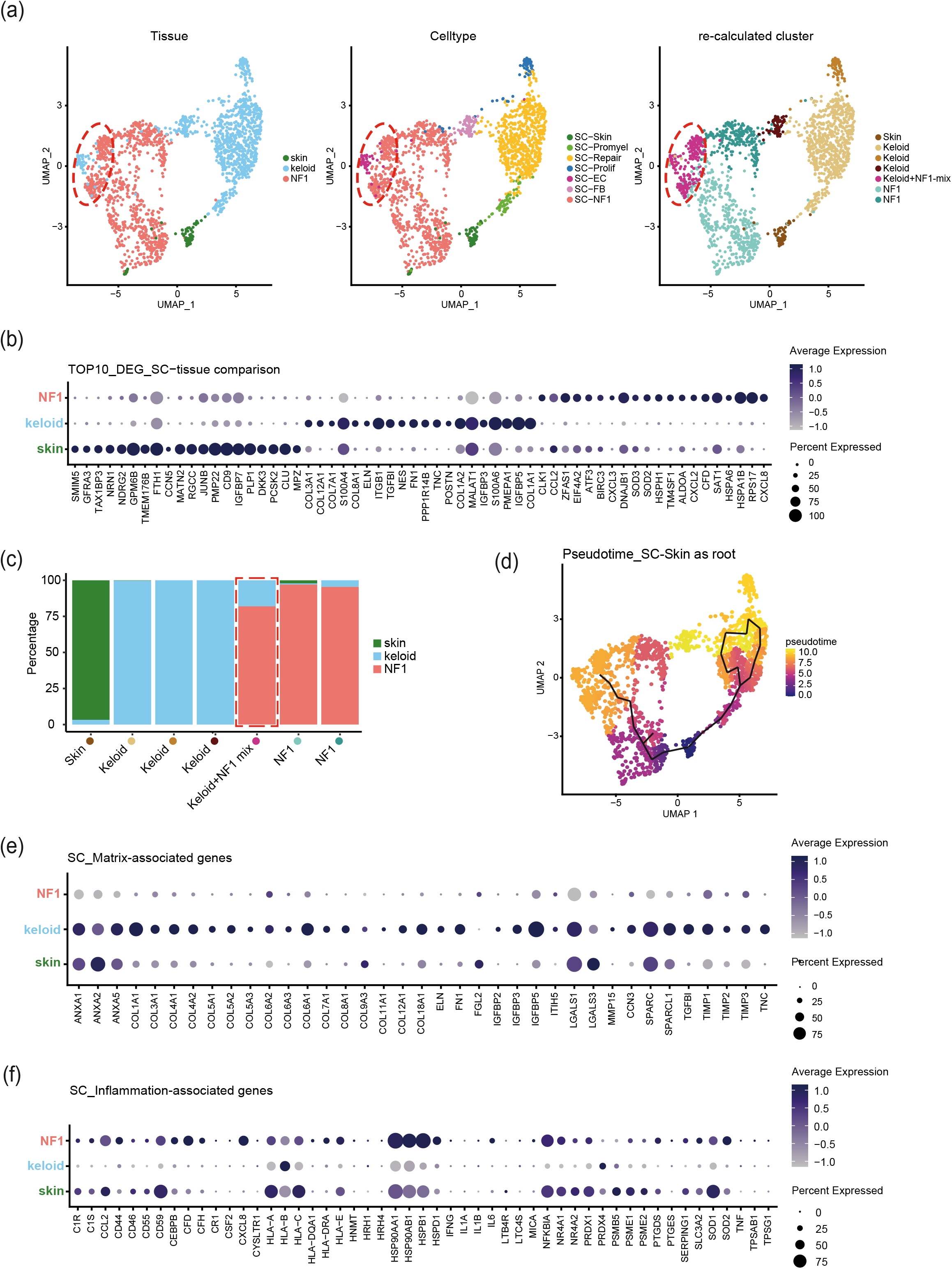
Keloidal Schwann cells differ from Schwann cells in cutaneous neurofibroma type 1. (a) UMAP-Plot of integrated Schwann cells data from healthy skin (n=7), keloid (n=4) and cutaneous neurofibroma type 1 (NF1) (n=3). Tissue UMAP-Plot indicates tissue-specific origin of cells. Celltype UMAP-Plot shows previously identified celltypes and NF1-derived cells. Re-calculated cluster UMAP-Plot coloured by newly formed cell cluster resulting from joint computation of all Schwann cells. Newly calculated clusters are titled based on their origin except the resulting mixed cluster (Keloid+NF1-mix) and marked by specific colour. Dashed circle highlights mixed cluster. (b) Top 10 differentially expressed genes between Schwann cells of healthy skin, keloid and NF1. Colours indicate average expression and circle indicate relative amounts of positive cells. (c) Percentage distribution of newly calculated clusters by tissue. Red frame highlights cluster with tissue purity below 80%. (d) UMAP-plot including pseudotime track starting from healthy skin Schwann cells (SC-Skin) to Schwann cells from Keloid or NF1. Colour code indicates cellular moment in the calculated pseudotime. (e) Expression of matrix-and (f) inflammation-associated genes by Schwann cells of healthy skin, keloids and NF1. Colours indicate average gene expression and size shows percentage of cells expressing the genes in a group.

### Schwann cells in keloids modulate macrophage function

Since denervated Schwann cells are known to interact with macrophages^26^, we next investigated the phenotype of macrophages and how macrophage function is influenced by the stroma in keloids, especially by Schwann cells. IF confirmed a reported increase of macrophages in keloids compared to healthy skin (Supplementary Figure 14a) ^33,34^. We therefore sub-clustered macrophages and classified them according to established M1 and M2 activation markers [macrophage mannose receptor (*MRC1*) and cluster of differentiation 163 (*CD163)* for M2-macrophages and interleukin-1 beta (*IL1B)* and chemokine ligand 2 (*CXCL2)* for M1-macrophages] (Figs. 8a). Whereas mainly M1-macrophages (MAC-M1; 89%) and only few M2-macrophages (MAC-M2; 8%) were detected in healthy skin, the majority of macrophages in keloids showed gene expression corresponding to M2-macrophages (MAC-M2; 49%). Another large macrophage population in keloids showed a mixed M1/M2 phenotype (MAC-M1/M2; 36%) and a smaller population was characterized by a combined expression of fibroblast- and macrophage-specific genes (MAC-FB; 15%) (Fig. 8b and Supplementary Fig. 14b).

**Figure 8.**
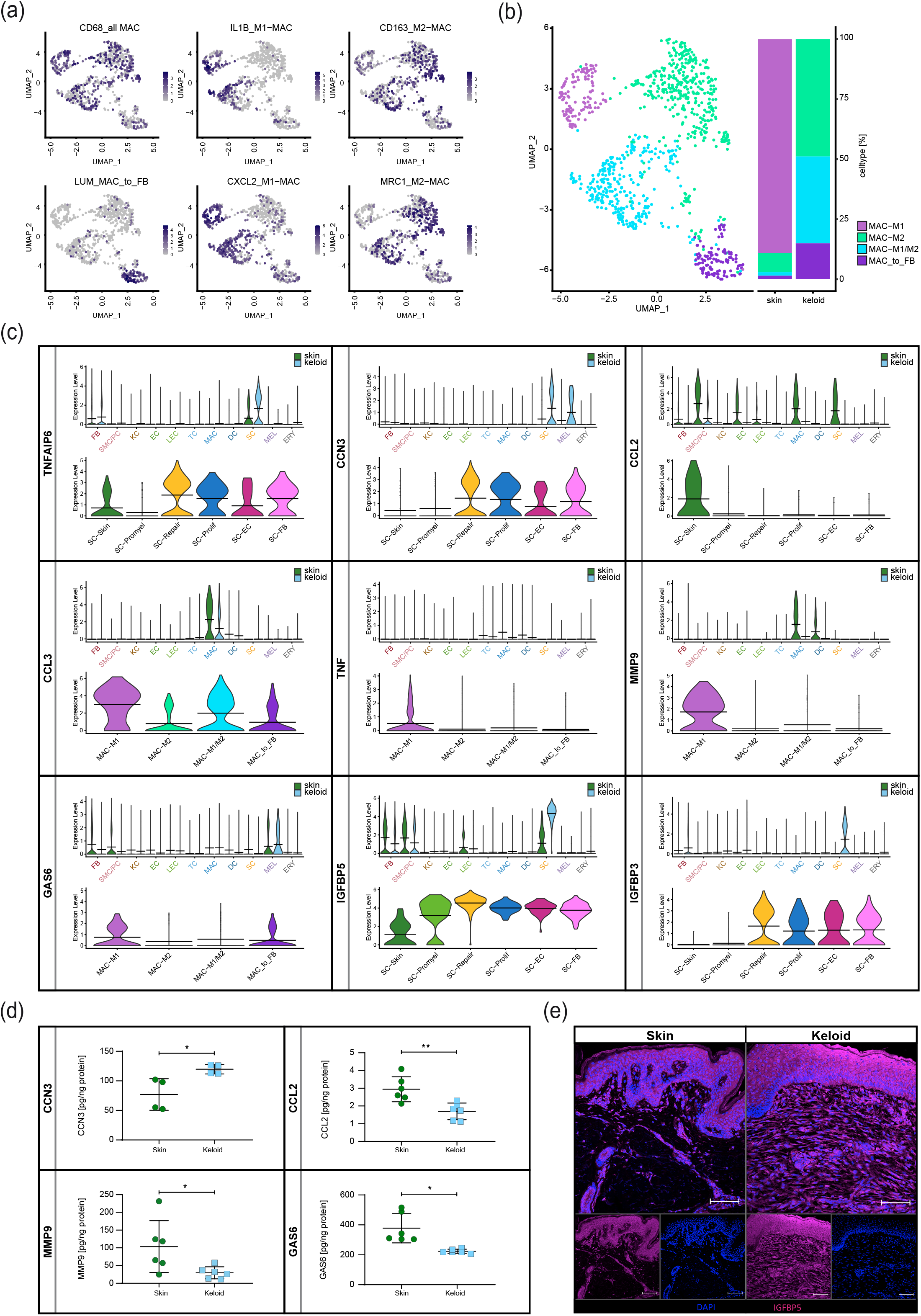
Predicted Schwann cell - Macrophage interaction in keloids. (a) Macrophage cluster identification by marker genes: cluster of differentiation 68 (*CD68*), interleukin 1 beta (*IL1B*), cluster of differentiation 163 (*CD163*), lumican (*LUM*), chemokine (c-x-c motif) ligand 2 (*CXCL2*), mannose receptor (*MRC1*). Colour intensity shows average gene expression. (b) Subset of Macrophages detected in healthy skin (n=4) and keloids (n=4). Bars depict percentage distribution of detected macrophage-cluster for each tissue. Identified macrophage cluster: M1-macrophages (MAC-M1), M2-macrophages (MAC-M2), M1-M2 intermediate macrophages (MAC-M1/M2), cells expressing macrophage and fibroblast specific genes (MAC_to_FB) (c) Expression of TNF-alpha induced protein 6 (*TNFAIP6)*, cellular communication network factor 3 (*CCN3*), CC-chemokine ligand 2 (*CCL2*), CC-chemokine ligand 3 (*CCL3*), TNF-alpha (*TNF*), matrix metallopeptidase 9 (*MMP9*), Growth arrest – specific 6 (*GAS6*), insulin-like growth factor-binding protein 3 (*IGFBP3*) and insulin-like growth factor-binding protein 5 (*IGFBP5*) of cells from healthy skin and keloids. Upper panel violin-plots depict gene expression across all celltypes split by tissue; green= skin, blue= keloid; fibroblasts (FB), smooth muscle cells and pericytes (SMC/PC), keratinocytes (KC), endothelial cells (EC), lymphatic endothelial cells (LEC), T-cells (TC), macrophages (MAC), dendritic cells (DC), Schwann cells (SC), melanocytes (MEL) and erythrocytes (ERY); Lower panel violin-plots show gene expression in previously described Schwann cell- or Macrophage-types; crossbeam marks mean gene expression; vertical lines show maximum expression, Width represents frequency of cells at the respective expression level. (d) Protein quantifications of CCN3, CCL2, MMP9, and GAS6 in skin and keloids (n = 2 donors for CCN3, n = 3 for all others). Whiskers represent first and third quartiles and central bar marks median. *p<0.05, **p<0.01; (e) Representative immunostaining of IGFBP5 in skin and keloids. Tissues of n = 3 donors per condition were stained. Scale bars: 100 µm.

We then focused our analysis on Schwann cell-derived factors that are known to influence macrophage function. Expression data of keloidal Schwann cells revealed several genes coding for secreted proteins, which are involved in the regulation of macrophage function. One of the strongest upregulated genes in SC-Repair was tumor necrosis factor alpha-induced protein 6 (*TNFAIP-6*) (Fig. 8c), which has been shown to inhibit inflammation^59-61^ and promote M2 polarization of macrophages^62,63^. In addition, we found an upregulation of *CCN3* in keloidal Schwann cells (Fig. 8c), a growth factor known to promote macrophage recruitment and differentiation into a M2 phenotype^64^. Interestingly, *CCN3* was also upregulated in melanocytes in keloids. In accordance with a M2-promoting environment, CC-chemokine ligand 2 (*CCL2*), a chemokine important for the recruitment of inflammatory M1 macrophages in wounds^65,66^, was almost completely absent in keloids (Fig. 8c). Furthermore, *CCL3* expression was strongly downregulated in keloids (Fig. 8c). Since CCL2 and CCL3 in combination with TNF-alpha (*TNF*) are known to enhance the production and release of the ECM-degrading enzyme matrix metallopeptidase 9 (MMP9) in monocytes^67^, we next investigated MMP9 levels in our data set. While *MMP9* was expressed by macrophages and dendritic cells in healthy skin, we detected no *MMP9* expression in keloids (Fig. 8c), and the total release of MMP9 protein was strongly reduced in keloids (Fig. 8d). Macrophages have been reported to regulate Schwann cell dynamics during nerve regeneration by supporting Schwann cell re-myelinisation and maturation through growth arrest-specific 6 (*GAS6*) ^68^. Indeed, *GAS6* expression was significantly decreased in MAC-M2 and MAC-M1/M2 in keloids (Fig. 8c). Among the strongest upregulated genes in SC-Repair was *IGFBP5*, a known pro-fibrotic factor supporting macrophage migration and conversion of monocytes into mesenchymal cells (Fig. 8c)^69-71^. IGFBP3, another member of the IGFBP protein family with known anti–inflammatory activity^72^ was also significantly upregulated in keloidal Schwann cells (Fig. 8c). To validate our scRNAseq data, we quantified several of the identified factors in skin and keloid biopsy lysates. We detected elevated CCN3 levels, while CCL2, MMP9, and GAS6 were significantly decreased in keloids compared to healthy skin (Fig. 8d). In addition, immunofluorescence staining confirmed increased IGFBP5 in the dermis of keloids compared to normal skin (Fig. 8e). Together, these data indicate a crosstalk of keloidal Schwann cells and macrophages promoting increased matrix production by Schwann cells whilst inhibiting matrix degradation by macrophages.

## Discussion

To date, the pathophysiological processes underlying keloid development are still poorly understood. Thus, we here performed a comprehensive scRNAseq approach of keloids supported by confocal microscopy imaging. Our findings introduce Schwann cells as novel players involved in keloid pathology, and support their plasticity as promising therapeutic target.

Unbiased clustering of our scRNAseq data confirmed high amounts of fibroblasts and endothelial cells in keloids compared to normal skin. This finding is in line with the known keloid morphology and a recently published single cell data set of keloids on gene regulation in these cell types^38^. In contrast to the publication by Liu *et al*., we detected a significantly increased Schwann cell population in keloids compared to healthy skin. Strikingly, the data set by Liu *et al*. showed a much higher Schwann cell number in the control skin samples as compared to our healthy skin samples. We explain this discrepancy by the different sources of skin control samples. While Liu *et al*., used samples from skin areas closely adjacent to the keloids^38^, we used normal skin from donors not harboring keloids. This observation indicates that the number of Schwann cells is already increased in skin adjacent to keloids and, therefore, already affected by keloid pathology. Since keloids are constantly growing tumors, infiltration of Schwann cells into the surrounding tissue might represent an important driver of the disease. However, further studies are necessary to address this question.

Tumor innervation and the contribution of the nervous system to keloid pathology have been hardly explored so far, and previous publications are contradictory^13-16^. As the major constituents of nerves are axons and Schwann cells, it was striking that the majority of keloidal Schwann cells were not associated with an axon. These axon-free Schwann cells had a spindle shaped morphology, comparable to that recently described for repair Schwann cells^20,24^. Repair Schwann cells show a characteristic expression pattern, including the expression of *JUN, SOX10, NGFR* and *NES*^21,39-42^, which we also verified in our keloidal Schwann cell cluster. Of note, some previously published markers for repair Schwann cells (*OLIG1, BDNF* of *GDNF*) were not detected in our data set, which might be due to higher detection limits in single cell sequencing analyses compared to bulk sequencing^20^. Alternatively, the long lasting presence of repair Schwann cells in keloids might affect gene expression, leading to the expression of a keloid-specific gene set differing from that of traditional repair Schwann cells. Repair Schwann cells are found in close proximity to the damaged/degrading axons, ultimately facilitating nerve regeneration^23^. Interestingly, Parfejevs *et al*. recently showed that peripheral glia cells are able to disseminate from the injured nerves into the granulation tissue, de-differentiate and proliferate during wound healing in mice^36^. At the wound site, infiltrating glia cells produce paracrine factors inducing myofibroblast differentiation, thereby promoting wound closure^36^. Of note, a connection between Schwann cell-density in the skin and impaired wound healing has also been demonstrated in humans^73^. Reinisch and co-workers showed that the number of Schwann cells in the periphery was strongly diminished in patients with diabetes mellitus, suggesting a role for Schwann cells in the development of diabetic foot ulcers^73^. The data obtained from mouse wounds^36^ are indeed in line with our Schwann cell data set of human keloids. Our pseudotime trajectory revealed that keloidal Schwann cells descend from de-differentiated adult Schwann cells and have the ability to gain a proliferative state and adapt characteristics of fibroblast-like cells or endothelial-like cells. This finding suggests that keloidal Schwann cells possess a long-lasting, highly plastic cell state. Importantly, these Schwann cells do not remain in normal scar tissue after completion of wound healing (Supplementary Fig. 15), further supporting our hypothesis that the persistence of repair Schwann cells in the skin significantly contribute to the pathogenesis of keloids. However, the exact mechanisms underlying this phenomenon remain unclear and need further investigations. Since mice do not develop keloids^1^ and currently available *in vitro* models have several limitations^1^, such as the lack of important cell types, including Schwann cells, answering this question is challenging and will require the establishment of new model systems. Nevertheless, we could demonstrate that Schwann cells significantly contribute to the stromal microenvironment of keloids, by expressing factors that directly affect the function of other cells and ECM formation (several collagens^56^ and members of the IGFBP-family)^56,69-71^.

The expression pattern of keloidal repair Schwann cells seems to demonstrate high disease-specificity, as Schwann cells of NF1, another benign skin tumor with Schwann cell contribution^57^, showed only minor similarities with those found in keloids. Whereas keloidal Schwann cells displayed a pro-fibrotic expression pattern with little expression of pro-inflammatory genes, Schwann cells in NF1 had a prominent pro-inflammatory phenotype, which is in line with a recent publication^74^. Although also in NF1, Schwann cells are not always attached to an axon, they do not develop a repair phenotype comparable to keloidal Schwann cells. This could be explained by the neoplastic transformation of Schwann cells during NF1 development ^75^, while the keloidal Schwann cells rather represent a deregulated repair cell persisting after an injury. Since a genetic component has been suggested for the development of keloids ^2,6-8^, it is conceivable that alterations of one or more genes might directly or indirectly affect Schwann cell functions. A recent study demonstrated that Schwann cells within the acute injury site or the denervated distal nerve segments adapt a different cellular behavior ^35^. Hence, the immediate and extended microenvironment of the wound and/or the forming scar is likely to be an important determinant for the unique cellular state of keloidal Schwann cells.

Our study revealed an important cross-talk between Schwann cells and macrophages (Supplementary Fig. 16). Macrophages are known to significantly contribute to the stromal milieu by affecting repair processes, tissue inflammation and matrix remodeling in a macrophage subtype-specific manner^30-32^. Whereas M1-macrophages are pro-inflammatory and matrix-degrading, M2-macrophages contribute to the composition of the ECM^31^. In line with previous publications^33,34^, we mainly detected M2-macrophages in our keloids. In addition, we identified one macrophage population with intermediate M1/M2 gene expression pattern and one subpopulation sharing gene sets specific for M2-macrophages and fibroblasts. The ability of macrophages to convert into mesenchymal cells is well documented and IGFBP5, one of the strongest expressed factors in keloidal Schwann cells, has been shown to support this process^71,76^. Although several genes associated with epithelial to mesenchymal transition, such as *SNAI1, SNAI2, ZEB1, ZEB2, TWIST1*, were not detected in this cell population (data not shown)^77^, our data indicate that, similar to Schwann cells, macrophages also show a high plasticity in keloids. Our data further revealed that repair Schwann cells present in keloids produced several factors affecting macrophage function. For example, we detected high levels of *TNFAIP6, IGFBP5* and *CCN3*, all known to regulate migration, activation and polarization of macrophages^62-64,71^. Interestingly, *CCL2* was strongly down-regulated in keloidal Schwann cells, contributing to significantly reduced overall CCL2 protein levels of keloids. As CCL2 is one of the most important factors provoking the accumulation of M1-type macrophages in the wound area^65,66^, reduced CCL2 levels might represent a crucial initial step in the development of keloids. *CCL3* and *TNF-α* were also down-regulated in keloids. All three factors together are known to be important for the production of macrophage-derived MMP9^67^. Indeed, MMP9 protein production was almost completely abolished in keloids. Interestingly, lack of MMP9 does not only contribute to less degradation of the ECM, but also affects Schwann cell function, as MMP9 has been shown to inhibit Schwann cell de-differentiation and proliferation^78^.

In summary, our study indicates that the interaction of repair-like Schwann cells and macrophages in keloids leads to increased matrix deposition, which could be responsible for their infinite growth. It is tempting to speculate that intervention at any point of this cycle might represent a promising treatment option. Our data suggest that especially the cellular plasticity of keloidal Schwann cells presents a promising therapeutic target. Unfortunately, little effort has been put in the development of proper model systems to study keloid formation and progression, and the few available models lack important cell types, including Schwann cells. Therefore, possible therapeutic options still need to be tested in costly *ex vivo* cultures or directly in clinical studies. This highlights the urgent need for standardized, high quality preclinical model systems to develop efficient treatment procedures for keloids. Together, our study opens a new perspective on the pathogenesis of keloids, which could significantly improve the treatment of this skin disease in the future.

## Methods

### Ethical statement

The use of resected skin and keloid tissue has been approved by the ethics committee of the Medical University of Vienna (votes 217/2010 and 1190/2020) in accordance with the guidelines of the Council for International Organizations of Medical Sciences (CIOMS). Written informed consent was obtained from all donors.

### Sample acquisition

ScRNAseq data of skin and neurofibroma were publicly available^37,58^. One additional skin sample was obtained from surplus abdominal skin after elective abdominoplasty. Keloid tissue samples were obtained from the earlobe (3 samples) and the chest (1 sample) after elective therapeutic resection. (donor information – Supplementary Table S1) Previous treatment of the area of interest by laser and/or radiation were defined as exclusion criteria. Diagnosis and surgical procedure were performed by plastic surgeons at the Department of Plastic, Reconstructive and Aesthetic Surgery of the Vienna General Hospital (Vienna, Austria).

### Sample dissociation and preparation of the single cell suspension

Tissue samples were washed with sterile Dulbecco’s phosphate-buffered saline (PBS, without Ca^2+^ and Mg^2+^, Gibco, Thermo Fisher Scientific, Waltham, MA, USA) under laminar air flow. Six mm punch biopsies were taken from the intact centre of the keloid and skin tissues, respectively. Biopsies were mechanically minced and enzymatically dissociated in gentleMACS C-Tubes (Miltenyi Biotec, Bergisch Gladbach, Germany) for 2.5 h at 37°C using MACS Miltenyi Whole Skin Dissociation Kit (Miltenyi,). Samples were further processed by gentleMACS OctoDissociator (Miltenyi,) according to the manufacturer’s protocol. Cell suspensions were sequentially passed through 100 µm and 40 µm cell strainer and washed twice with 0.04 % bovine serum albumin (BSA, Sigma Aldrich, St. Louis, MO, USA) in PBS. Cell concentrations and viability were assessed by Acridine Orange/Propidium Iodide (AO/PI) Cell Viability Kit (Logos Biosystems, Anyang-si, Gyeonggi-do, South Korea) and detected by LUNA-FL™ Dual Fluorescence Cell Counter (Logos Biosystems). In case of cell viability <80%, the samples were treated with Dead Cell Removal Kit (Miltenyi). Cell concentrations were adjusted to 0.7-1.2⨯10^6^ cells/ml. Samples were kept on ice until further processing. Gel beads-in-emulsion (GEMs) were generated within 4 hours after tissue resection.

### Single cell processing and library preparation

GEM generation, barcoding, sample clean-up, cDNA amplification, and library construction were performed according to manufacturer’s protocol using Chromium Next GEM Single Cell 3’GEM, Library & Gel Bead Kit v3.1, Chromium Next GEM Chip G Single Cell Kit, and Single Index Kit T Set A (all 10x Genomics, Pleasanton, CA, USA).

### Sequencing and matrix preparation

RNA-sequencing, demultiplexing, and counting were carried out by the Biomedical Sequencing Facility (BSF) of the Center for Molecular Medicine (CeMM, Vienna, Austria). Samples were sequenced paired end with dual indexing (read length 75bp) in foursome pools using a HiSeq 3000/4000 (Illumina, San Diego, CA, USA). Raw data were aligned to the human reference genome (GRCh38) and counted by the Cellranger pipelines (Cellranger v3.0.2, except skin 7 v5.0.1, 10x Genomics).

### Bioinformatical analysis

Bioinformatics analyses were performed using R (R v4.0.3, The R Foundation, Vienna, Austria), R-studio and Seurat (Seurat v4.0.0, Satija Lab)^79^.

All included data sets were aligned by features. Only features detected in all datasets have been included, feature doublets have been excluded and feature names unified. Data were pre-processed by sctransform-normalization supported by the glmGamPoi package and integrated according to Seurat Vignette^80,81^. PCA and UMAP were calculated. All subset analyses have been performed as new calculation based on the raw data of the cells of interest. For cell type identification, clustermarker features were calculated and well-established marker genes were chosen to verify the assignment (marker genes information – Supplementary Table S2). Pseudotime-trajectory calculation was performed using Monocle3 (Monocle3, v.0.2.3.0, Trapnell Lab)^82-86^. The S4 objects of class Seurat were converted to a cell_data_set_object keeping the generated UMAP. Thereby, the principal graphs were calculated based on the cell distribution generated in Seurat. The correlations of the transcriptomic expression pattern of all detected Schwann cells were high enough to form one partition. A total of 20 graph centres (skin-keloid) and 40 graph centres (skin-keloid-NF1) was determined. The Schwann cell cluster of skin samples was defined as the root for pseudotime calculations. Expression changes along the trajectory were calculated for Schwann cells from intact skin to keloids. Gene ontology (GO) enrichment analysis was performed based on gene lists of clustermarker calculations and differentially expressed gene calculations. Only genes above avg_logFC of 2 were included. In case of less than 15 or more than 100 genes with an avg_logFC cut-off of 2 or higher, an avg_logFC-threshold of 1.5 and 3 was set, respectively. For enrichment analysis, Metascape was used^87^. A p-value cutoff of 0.05 and a minimum enrichment score of 2 was set.

### Immunofluorescence

For cryopreservation, tissues were washed with PBS and fixed in 4.5% formaldehyde solution, neutral buffered (SAV Liquid Production GmbH, Flintsbach am Inn, Germany) for 24 hours at 4°C. Specimens were washed with PBS for 24 hours and dehydrated by sequential incubation with 10%, 25%, and 42% succrose for 24 hours each. Tissues were snap-frozen in optimal cutting temperature compound (OCT compound, TissueTek, Sakura, Alphen aan den Rijn, The Netherlands) and stored at −80°C. Ten µm sections were cut using a cryotome (Leica, Wetzlar, Germany) and dried for 30 min at room temperature. Cryosections were immersed in PBS followed by blocking and permeabilization with 1% BSA, 5% goat serum (DAKO, Glostrup, Denmark), and 0.3% Triton-X (Sigma Aldrich) in PBS for 15 min.

For paraffin embedding, tissues were washed with PBS and 6 mm punches were obtained. Biopsies were cut in a half and each part was fixed in 4.5% formaldehyde solution overnight and embedded in paraffin. After de-paraffinization and hydration, sections were boiled in Target Retrieval Solution (DAKO) using a 2100 Antigen Retriever (DAKO) followed by three washes with PBS for 5 minutes.

Antibody details, dilutions, and incubation times are listed in Supplementary Table S3. A washing step consisted of three washes with PBS for 5 minutes and was performed after each antibody incubation step.

If stained for S100 (DAKO), primary antibodies were diluted in the ready to use S100 antibody solution. For the other stainings, antibodies were diluted in the antibody staining solution containing 1% BSA and 0.1 % Triton-X in PBS. Sections were incubated with secondary antibodies for 1 hour and nuclear stain was performed by adding 50 µg/mL 4,6-diamidino-2-phenylindole (DAPI, Thermo Fisher Scientific) in PBS for 2 minutes. Sections were embedded in mounting medium (Fluoromount-G, SouthernBiotech, Birmingham, AL, USA) and stored at 4°C. Micrographs were acquired with a confocal laser scanning microscope (TCS SP8X, Leica) equipped with a 10x (0.3 HCPL FluoTar), a 20x (0.75 HC-Plan-Apochromat, Multimmersion), a 20x (0.75 HC-Plan-Apochromat) and a 63x (1.3 HC-Plan-Apochromat, Glycerol) objective using Leica application suite X version 1.8.1.13759 or LAS AF Lite software (both Leica). Confocal images are depicted as maximum projection of total z-stacks. All stainings were performed on cryo-as well as paraffin-preserved tissue samples.

### Enzyme-linked immunosorbent assay (ELISA)

Skin- and keloid-derived proteins were quantified by ELISA. Six mm punch biopsies were collected and cryopreserved at −80 °C until further processing. Tissues were lysed in 0.5 ml of 0.1% Triton X in PBS and homogenised (Precellys 24 Homogenisator, Bertin Instruments, Montigny-le-Bretonneux, France). Homogenization was repeated after overnight incubation at 4°C. Sample were centrifuged for 15 min at 13,000 *g* and supernatants were used for protein quantifications. Total protein amounts were determined using Micro BCA^™^ Protein Assay Kit (Thermo Fisher Scientific) according to manufacturer’s protocol. CCL2, MMP9, CCN3, and GAS6 concentrations were determined using commercially available ELISA kits as recommended by the manufacturer (Human CCL2/MCP-1 Quantikine ELISA Kit, Human MMP-9 DuoSet ELISA Kit, Human NOV/CCN3 DuoSet ELISA Kit, and Human GAS6 DuoSet ELISA Kit (all R&D Systems, Minneapolis, MN, USA).

### Statistical analyses

For statistical evaluation, GraphPad Prism 8 software (GraphPad Software Inc., La Jolla, CA, USA) was used. Normal distribution within a group was tested by Shapiro-Wilk test. Comparison between two groups with normal distribution was performed with paired t-test. Independent groups without normal distribution were compared by Mann-Whitney-U-Test. Asterisks were used to mark p-values: *p<0.05, **p<0.01, ***p<0.001, ****p<0.0001.

### Data availability

ScRNASeq data are available in NCBI’s Gene Expression Omnibus (GEO) and accessible through GEP series accession number GSE181316.

## Data Availability

ScRNASeq data will be available after publication upon request.

## Conflict of interest

The authors declare that the research has been performed without any conflict of interest.

## Acknowledgment

The present study was financed by the FFG Grant “APOSEC” (852748 and 862068; 2015-2019), the Vienna Business Agency “APOSEC to clinic” (ID 2343727, 2018-2020) and by the Aposcience AG under the direction of group leader HJA. The Sparkling Science Program of the Austrian Federal Ministry of Education, Science and Research (SPA06/055) funded MM. We would like to thank Hans Peter Haselsteiner and the CRISCAR Familienstiftung for their ongoing support of the Medical University/Aposcience AG public private partnership aiming to augment basic and translational clinical research in Austria/Europe. The authors acknowledge the core facilities of the Medical University of Vienna, a member of Vienna Life Science Instruments. We also thank Matthias Wielscher for his support in bioinformatics issues. We thank Barbara Messner and her team for providing the SMA antibody used in this study.

## Author contributions

MD, TW, ET, HJA and MM provided study design and concept; VV, WH and CR provided patient sample material; HJA and MM acquired funding; MD, TW, DC, CM, KK, DB, MF, LS and BG prepared samples and conducted staining and experiments. MD, TW, ML and MM performed data analysis, visualization and figure design; MD, TW, ET and MM participated in data interpretation; MD, ML and MM drafted the manuscript.

All Authors reviewed the manuscript.

## Supplementary Figure Legends

Supplementary Fig.1

<u>Celltype identification by marker gene expression in cells of healthy skin and keloids</u>

(a) Feature Plots of well-known marker genes: lumican (*LUM*), decorin (*DCN*), fibulin-1 (*FBLN1*) for fibroblasts, actin alpha 2 (*ACTA2*) for smooth muscle cells, regulator of g-protein signalling 5 (*RGS5*) for pericytes, keratin 1 (*KRT1*), keratin 14 (*KRT14*) for keratinocytes, intercellular adhesion molecule 1 (*ICAM1*) for endothelial cells, lymphatic vessel endothelial hyaluronan receptor 1 (*LYVE1*) for lymphatic endothelial cells, cluster of differentiation 3D (*CD3D*) for T cells, cluster of differentiation 68 (*CD68*) for macrophages, FC fragment of IgE receptor Ia (*FCER1A*) for dendritic cells, S100 calcium binding protein B (*S100B*) for Schwann cells, melan-A (*MLANA*) for melanocytes, hemoglobin subunit beta (*HBB*) for erythrocytes. In Feature Plots the normal log expression of the respective gene is mapped on the UMAP-Plot. (b) Dotplot of marker genes: platelet-derived growth factor receptor A (*PDGFRA*), LUM, collagen type I alpha 1 (*COL1A1*), *DCN, FBLN1* for fibroblasts (FB), *ACTA2, RGS5* for smooth muscle cells and pericytes (SMC/PC), keratin 10 (*KRT10*), *KRT1, KRT14*, keratin 5 (*KRT5*) for keratinocytes (KC), E-selectin (*SELE*), von willebrand factor (*VWF*) for endothelial cells (EC), *LYVE1* for lymphatic endothelial cells (LEC), *CD3D*, cluster of differentiation 2 (*CD2*), c-x-c chemokine receptor type 4 (*CXCR4*) for T-cells (TC), *CD68*, allograft inflammatory factor 1 (*AIF1*) for macrophages (MAC), *FCER1A* for dendritic cells (DC), *S100B*, nerve growth factor receptor (*NGFR*) for Schwann cells (SC), premelanosome protein (*PMEL*), *MLANA* for melanocytes (MEL), *HBB*, haemoglobin subunit alpha 1 (*HBA1*) for erythrocytes (ERY). Colour code indicates log gene expressions.

Supplementary Fig.2

<u>Tissue comparison of healthy skin and keloids</u>

(a) Representative Hematoxylin-Eosin-staining of skin and keloid sections. Representative immunostainings of (b) smooth muscle alpha-actin (SMA) and (c) vimentin (VIM) in healthy skin compared to keloid tissue. Tissues of n = 3 donors per condition were stained. Scale bars: 100 µm.

Supplementary Fig.3

<u>Schwann cell-subtypes distinguish between healthy skin and keloids</u>

(a) UMAP-Plot of Schwann cell subtypes. Bars display absolute numbers of the identified cell subtypes in skin and keloid. Identified Schwann cell cluster: Myelinating and non-myelinating Schwann cells of the healthy skin (SC-Skin), promyelinating Schwann cells (SC-Promyel), repair Schwann cells (SC-Repair), proliferating Schwann cells (SC-Prolif), cells expressing Schwann cell and endothelial cell specific genes (SC-EC), cells expressing Schwann cell and fibroblast specific genes (SC-FB).

Supplementary Fig. 4

<u>SC-EC represents an intermediate cellular state between Schwann cells and endothelial cells</u>

(a) Total number of differentially expressed genes with average log foldchange ≥ 2 comparing SC-EC with all remaining Schwann cells (SC-Skin, SC-Promyel, SC-Repair, SC-Prolif, SC-FB). To determine DEGs, either endothelial and lymphatic endothelial cells (EC + LEC) or keratinocytes were used as a reference. Green marks upregulation, red marks downregulation, top 10 upregulated genes are listed. (b) Feature Plot of E-selectin (*SELE*) (red) and S100 calcium binding protein B (*S100B*) (green) expression of Schwann cells from skin and keloid as well as feature-blend revealing doublepositive cells in yellow.

Supplementary Fig. 5

<u>SC-FB represents an intermediate cellular state between Schwann cells and fibroblasts</u>

(a) Comparison of SC-FB with all remaining Schwann cells (SC-Skin, SC-Promyel, SC-Repair, SC-Prolif, SC-EC), with all fibroblasts or with all keratinocytes as a reference. Bar diagrams depict total number of differentially expressed genes with average logfoldchange ≥ 2. Green = UP-regulation, red = Down-regulation, Top10 upregulated genes for the corresponding comparison are shown. (b) Feature Plot blend of cluster of differentiation 90 (*THY1*) and S100B, red= THY1 expression, green= S100B expression, yellow = double positive cells.

Supplementary Fig. 6

<u>Transcriptional changes governing the transition from mature skin Schwann cells to repair Schwann cells</u>

Pseudotime Plots show individual changes in gene expression along the pseudotime of the transition from mature skin Schwann cells to repair Schwann cells.

Supplementary Fig. 7

<u>Skin-Schwann cells reveal characteristic functions by GO-term analysis</u>

(a) GO-term analysis of top-clustermarker of SC-skin. (b) Enrichment analysis of SC-Promyel using top-clustermarker with foldchange ≥ 2. Statisticial significance of terms is sympbolized by bar length. Red boxes in Gridplot mark annotated genes. Dotplot shows expression of top-clustermarker by the different Schwann cell types. Colour indicates average gene expression. Dot-size shows the percentage of cells in a group expressing the gene.

Supplementary Fig. 8

<u>GO-term analysis confirms Schwann cell subtype characterisation</u>

Enrichment analysis of top-clustermarker genes with average log-foldchange ≥ 2 of SC-Prolif, SC-EC and SC-FB. Bars depict statistical significance; red boxes mark annotated genes; Dots show expression of genes in the different Schwann cell subtypes. Size of dots symbolizes percentage of cells in a group expressing the gene. Dot colour shows average gene expression levels.

Supplementary Fig. 9

<u>Expression of connective tissue-associated genes in cells of healthy skin and keloids</u>

(a) Comparing the expression of matrix-associated genes in all detected cells of skin vs. all keloid cells. Crossbeam mark mean value; ns p-value >0.05, ***p-value <0.001, ****p-value <0.0001; vertical lines show maximum expression, Width represents frequency of cells at the respective gene expression level. cellular communication network factor 3 (*CCN3*), collagen type I alpha 1 (*COL1A1*), collagen type III alpha 1 (*COL3A1*), collagen type IV alpha 1 (*COL4A1*), collagen type IV alpha 2 (*COL4A2*), collagen type V alpha 1 (*COL5A1*), collagen type V alpha 2(*COL5A2*), collagen type VII alpha 1 (*COL7A1*), collagen type VIII alpha 1 (*COL8A1*), collagen type XII alpha 1 (*COL12A1*), collagen type XVIII alpha 1 (*COL18A1*), elastin (*ELN*), insulin like growth factor binding protein 3 (*IGFBP3*), insulin like growth factor binding protein 5 (*IGFBP5*), tenascin c (*TNC*), transforming growth factor beta induced (*TGFBI*). Expression-comparison of (b) glycoproteins, (c) proteoglycans and (d) collagens between the cells of healthy skin and keloids. fibroblasts (FB), smooth muscle cells and pericytes (SMC/PC), keratinocytes (KC), endothelial cells (EC), lymphatic endothelial cells (LEC), T-cells (TC), macrophages (MAC), dendritic cells (DC), Schwann cells (SC), melanocytes (MEL) and erythrocytes (ERY); Dot-size depicts percentage of cells in a group expressing the gene. Dot-colour symbolizes average gene expression.

Supplementary Fig. 10

<u>Expression of ECM-associated genes in cells of healthy skin and keloids</u>

Comparison of ECM regulator (a) and ECM affiliated proteins (b) between the cells of healthy skin and keloids. fibroblasts (FB), smooth muscle cells and pericytes (SMC/PC), keratinocytes (KC), endothelial cells (EC), lymphatic endothelial cells (LEC), T-cells (TC), macrophages (MAC), dendritic cells (DC), Schwann cells (SC), melanocytes (MEL) and erythrocytes (ERY); Dot-size depicts percentage of cells in a group expressing the gene. Dot-colour symbolizes average gene expression.

Supplementary Fig. 11

<u>Expression of secreted factors in cells of healthy skin and keloids</u>

(a) Comparison of secreted factors between the cells of healthy skin and keloids. fibroblasts (FB), smooth muscle cells and pericytes (SMC/PC), keratinocytes (KC), endothelial cells (EC), lymphatic endothelial cells (LEC), T-cells (TC), macrophages (MAC), dendritic cells (DC), Schwann cells (SC), melanocytes (MEL) and erythrocytes (ERY); Dot-size depicts percentage of cells in a group expressing the gene. Dot-colour symbolizes average gene expression.

Supplementary Fig. 12

<u>Expression of immunological genes in cells of healthy skin and keloids</u>

(a) Comparing the expression of inflammation-associated genes in all detected cells of skin vs. all keloidal cells. Crossbeams mark mean value; *p-value <0.05, **p-value <0.01, ****p-value <0.0001; vertical lines show maximum expression, respective shape represents all results; respective width represents frequency of cells at the appropriate expression level. interleukin 1 alpha (*IL1A*), interleukin 1 beta (*IL1B*), interleukin 6 (*IL6*), interleukin 8 (*CXCL8*), tumor necrosis factor alpha (*TNF*), interferon gamma (*IFNG*), leukotriene C4 synthase (*LTC4S*) and colony-stimulating factor 2 (*CSF2*). (b) Expression-comparison of immunology associated genes between the cells of healthy skin and keloids. fibroblasts (FB), smooth muscle cells and pericytes (SMC/PC), keratinocytes (KC), endothelial cells (EC), lymphatic endothelial cells (LEC), T-cells (TC), macrophages (MAC), dendritic cells (DC), Schwann cells (SC), melanocytes (MEL) and erythrocytes (ERY); Dot-size depicts percentage of cells in a group expressing the gene. Dot-colour symbolizes average gene expression.

Supplementary Fig. 13

<u>GO-term analysis confirms similarities between SC-EC and new formed NF1-keloid mix cluster</u>

(a) Enrichment analysis of top-clustermarker genes with average log-foldchange ≥ 2 of “Keloid+NF1-mix” cluster. Bars depict statistical significance; red boxes mark term annotated genes; Dots show expression of genes in the corresponding new calculated clusters. Size of dots symbolizes percentage of cells in a group expressing the gene. Dot-colour symbolizes average gene expression.

Supplementary Fig. 14

<u>Increase of macrophages in keloids compared to healthy skin</u>

(a) Immunostainings of macrophages (CD68) and Schwann cells (S100B) in skin and keloid. Scale bars: 100 µm. (b) UMAP-Plot of macrophage subtypes split by tissue Identified macrophage cluster: M1-macrophages (MAC-M1), M2-macrophages (MAC-M2), M1-M2 intermediate macrophages (MAC-M1/M2), cells expressing macrophage and fibroblast specific genes (MAC_to_FB).

Supplementary Fig. 15

<u>Increase of macrophages in keloids compared to healthy skin</u>

Comparison of Schwann cell-percentages from skin (n=7), normal scar (n=3) and keloid (n=4).

Supplementary Fig. 16

<u>Graphical scheme of Schwann cells in healthy skin and keloids</u>

## Supplementary Table Legends

Supplementary Table 1 – donor information

---

Supplementary Table 2 – marker genes information

Supplementary Table 3 – antibody information

## Notes

### Competing Interest Statement

The authors have declared no competing interest.

### Author Declarations

The use of resected skin and keloid tissue has been approved by the ethics committee of the Medical University of Vienna (votes 217/2010 and 1190/2020) in accordance with the guidelines of the Council for International Organizations of Medical Sciences (CIOMS).

## References

1 Limandjaja, G. C., Niessen, F. B., Scheper, R. J. & Gibbs, S. The Keloid Disorder: Heterogeneity, Histopathology, Mechanisms and Models. Front Cell Dev Biol 8, 360, doi:10.3389/fcell.2020.00360 (2020).

2 English, R. S. & Shenefelt, P. D. Keloids and hypertrophic scars. Dermatol Surg 25, 631–638, doi:10.1046/j.1524-4725.1999.98257.x (1999).

3 Murray, J. C. Keloids and hypertrophic scars. Clin Dermatol 12, 27–37, doi:10.1016/0738-081x(94)90254-2 (1994).

4 Ud-Din, S. & Bayat, A. New insights on keloids, hypertrophic scars, and striae. Dermatol Clin 32, 193–209, doi:10.1016/j.det.2013.11.002 (2014).

5 Balci, D. D., Inandi, T., Dogramaci, C. A. & Celik, E. DLQI scores in patients with keloids and hypertrophic scars: a prospective case control study. J Dtsch Dermatol Ges 7, 688–692, doi:10.1111/j.1610-0387.2009.07034.x (2009).

6 Alhady, S. M. & Sivanantharajah, K. Keloids in various races. A review of 175 cases. Plast Reconstr Surg 44, 564–566, doi:10.1097/00006534-196912000-00006 (1969).

7 Ramakrishnan, K. M., Thomas, K. P. & Sundararajan, C. R. Study of 1,000 patients with keloids in South India. Plast Reconstr Surg 53, 276–280, doi:10.1097/00006534-197403000-00004 (1974).

8 Kiprono, S. K. et al. Epidemiology of keloids in normally pigmented Africans and African people with albinism: population-based cross-sectional survey. Br J Dermatol 173, 852–854, doi:10.1111/bjd.13826 (2015).

9 Mustoe, T. A. et al. International clinical recommendations on scar management. Plast Reconstr Surg 110, 560–571, doi:10.1097/00006534-200208000-00031 (2002).

10 Khansa, I., Harrison, B. & Janis, J. E. Evidence-Based Scar Management: How to Improve Results with Technique and Technology. Plast Reconstr Surg 138, 165s–178s, doi:10.1097/prs.0000000000002647 (2016).

11 Reinisch, C. M. & Tschachler, E. The dimensions and characteristics of the subepidermal nerve plexus in human skin--terminal Schwann cells constitute a substantial cell population within the superficial dermis. J Dermatol Sci 65, 162–169, doi:10.1016/j.jdermsci.2011.10.009 (2012).

12 Ashrafi, M., Baguneid, M. & Bayat, A. The Role of Neuromediators and Innervation in Cutaneous Wound Healing. Acta Derm Venereol 96, 587–594, doi:10.2340/00015555-2321 (2016).

13 Hochman, B. et al. Nerve fibres: a possible role in keloid pathogenesis. Br J Dermatol 158, 651–652, doi:10.1111/j.1365-2133.2007.08401.x (2008).

14 Saffari, T. M. et al. Sensory perception and nerve fibre innervation in patients with keloid scars: an investigative study. Eur J Dermatol 28, 828–829, doi:10.1684/ejd.2018.3405 (2018).

15 Drummond, P. D., Dawson, L. F., Wood, F. M. & Fear, M. W. Up-regulation of α(1)-adrenoceptors in burn and keloid scars. Burns 44, 582–588, doi:10.1016/j.burns.2017.09.010 (2018).

16 Shoukrey, N. M. & Tabbara, K. F. Ultrastructural study of a corneal keloid. Eye (Lond) 7 (Pt 3), 379–387, doi:10.1038/eye.1993.76 (1993).

17 Tschachler, E. et al. Sheet preparations expose the dermal nerve plexus of human skin and render the dermal nerve end organ accessible to extensive analysis. J Invest Dermatol 122, 177–182, doi:10.1046/j.0022-202X.2003.22102.x (2004).

18 Nave, K. A. & Trapp, B. D. Axon-glial signaling and the glial support of axon function. Annu Rev Neurosci 31, 535–561, doi:10.1146/annurev.neuro.30.051606.094309 (2008).

19 Riethmacher, D. et al. Severe neuropathies in mice with targeted mutations in the ErbB3 receptor. Nature 389, 725–730, doi:10.1038/39593 (1997).

20 Jessen, K. R. & Mirsky, R. The repair Schwann cell and its function in regenerating nerves. J Physiol 594, 3521–3531, doi:10.1113/jp270874 (2016).

21 Arthur-Farraj, P. J. et al. c-Jun reprograms Schwann cells of injured nerves to generate a repair cell essential for regeneration. Neuron 75, 633–647, doi:10.1016/j.neuron.2012.06.021 (2012).

22 Jessen, K. R., Mirsky, R. & Arthur-Farraj, P. The Role of Cell Plasticity in Tissue Repair: Adaptive Cellular Reprogramming. Dev Cell 34, 613–620, doi:10.1016/j.devcel.2015.09.005 (2015).

23 Jessen, K. R. & Mirsky, R. The Success and Failure of the Schwann Cell Response to Nerve Injury. Front Cell Neurosci 13, 33, doi:10.3389/fncel.2019.00033 (2019).

24 Gomez-Sanchez, J. A. et al. After Nerve Injury, Lineage Tracing Shows That Myelin and Remak Schwann Cells Elongate Extensively and Branch to Form Repair Schwann Cells, Which Shorten Radically on Remyelination. J Neurosci 37, 9086–9099, doi:10.1523/jneurosci.1453-17.2017 (2017).

25 Stoll, G. & Müller, H. W. Nerve injury, axonal degeneration and neural regeneration: basic insights. Brain Pathol 9, 313–325, doi:10.1111/j.1750-3639.1999.tb00229.x (1999).

26 Tofaris, G. K., Patterson, P. H., Jessen, K. R. & Mirsky, R. Denervated Schwann cells attract macrophages by secretion of leukemia inhibitory factor (LIF) and monocyte chemoattractant protein-1 in a process regulated by interleukin-6 and LIF. J Neurosci 22, 6696–6703, doi:10.1523/jneurosci.22-15-06696.2002 (2002).

27 Gomez-Sanchez, J. A. et al. Schwann cell autophagy, myelinophagy, initiates myelin clearance from injured nerves. J Cell Biol 210, 153–168, doi:10.1083/jcb.201503019 (2015).

28 Jang, S. Y. et al. Autophagic myelin destruction by Schwann cells during Wallerian degeneration and segmental demyelination. Glia 64, 730–742, doi:10.1002/glia.22957 (2016).

29 Yoshiba, N. et al. M2 Phenotype Macrophages Colocalize with Schwann Cells in Human Dental Pulp. J Dent Res 99, 329–338, doi:10.1177/0022034519894957 (2020).

30 Lucas, T. et al. Differential roles of macrophages in diverse phases of skin repair. J Immunol 184, 3964–3977, doi:10.4049/jimmunol.0903356 (2010).

31 Zhu, Z., Ding, J., Ma, Z., Iwashina, T. & Tredget, E. E. Systemic depletion of macrophages in the subacute phase of wound healing reduces hypertrophic scar formation. Wound Repair Regen 24, 644–656, doi:10.1111/wrr.12442 (2016).

32 Mirza, R., DiPietro, L. A. & Koh, T. J. Selective and specific macrophage ablation is detrimental to wound healing in mice. Am J Pathol 175, 2454–2462, doi:10.2353/ajpath.2009.090248 (2009).

33 Bagabir, R. et al. Site-specific immunophenotyping of keloid disease demonstrates immune upregulation and the presence of lymphoid aggregates. Br J Dermatol 167, 1053–1066, doi:10.1111/j.1365-2133.2012.11190.x (2012).

34 Boyce, D. E., Ciampolini, J., Ruge, F., Murison, M. S. & Harding, K. G. Inflammatory-cell subpopulations in keloid scars. Br J Plast Surg 54, 511–516, doi:10.1054/bjps.2001.3638 (2001).

35 Clements, M. P. et al. The Wound Microenvironment Reprograms Schwann Cells to Invasive Mesenchymal-like Cells to Drive Peripheral Nerve Regeneration. Neuron 96, 98-114.e117, doi:10.1016/j.neuron.2017.09.008 (2017).

36 Parfejevs, V. et al. Injury-activated glial cells promote wound healing of the adult skin in mice. Nat Commun 9, 236, doi:10.1038/s41467-017-01488-2 (2018).

37 Tabib, T., Morse, C., Wang, T., Chen, W. & Lafyatis, R. SFRP2/DPP4 and FMO1/LSP1 Define Major Fibroblast Populations in Human Skin. J Invest Dermatol 138, 802–810, doi:10.1016/j.jid.2017.09.045 (2018).

38 Liu, X. et al. Single-cell RNA-seq reveals lineage-specific regulatory changes of fibroblasts and vascular endothelial cells in keloids. J Invest Dermatol, doi:10.1016/j.jid.2021.06.010 (2021).

39 Mirsky, R. et al. Novel signals controlling embryonic Schwann cell development, myelination and dedifferentiation. J Peripher Nerv Syst 13, 122–135, doi:10.1111/j.1529-8027.2008.00168.x (2008).

40 Finzsch, M. et al. Sox10 is required for Schwann cell identity and progression beyond the immature Schwann cell stage. J Cell Biol 189, 701–712, doi:10.1083/jcb.200912142 (2010).

41 Britsch, S. et al. The transcription factor Sox10 is a key regulator of peripheral glial development. Genes Dev 15, 66–78, doi:10.1101/gad.186601 (2001).

42 Weiss, T. et al. Proteomics and transcriptomics of peripheral nerve tissue and cells unravel new aspects of the human Schwann cell repair phenotype. Glia 64, 2133–2153, doi:10.1002/glia.23045 (2016).

43 Ackerman, S. D., Garcia, C., Piao, X., Gutmann, D. H. & Monk, K. R. The adhesion GPCR Gpr56 regulates oligodendrocyte development via interactions with Gα12/13 and RhoA. Nat Commun 6, 6122, doi:10.1038/ncomms7122 (2015).

44 Barlow, J. Z. & Huntley, G. W. Developmentally regulated expression of Thy-1 in structures of the mouse sensory-motor system. J Comp Neurol 421, 215–233, doi:10.1002/(sici)1096-9861(20000529)421:2<215::aid-cne7>3.0.co;2-u (2000).

45 Quintes, S. et al. Zeb2 is essential for Schwann cell differentiation, myelination and nerve repair. Nat Neurosci 19, 1050–1059, doi:10.1038/nn.4321 (2016).

46 Salim, C., Boxberg, Y. V., Alterio, J., Féréol, S. & Nothias, F. The giant protein AHNAK involved in morphogenesis and laminin substrate adhesion of myelinating Schwann cells. Glia 57, 535–549, doi:10.1002/glia.20782 (2009).

47 Mikol, D. D., Scherer, S. S., Duckett, S. J., Hong, H. L. & Feldman, E. L. Schwann cell caveolin-1 expression increases during myelination and decreases after axotomy. Glia 38, 191–199, doi:10.1002/glia.10063 (2002).

48 Weiss, T. et al. Schwann cell plasticity regulates neuroblastic tumor cell differentiation via epidermal growth factor-like protein 8. Nat Commun 12, 1624, doi:10.1038/s41467-021-21859-0 (2021).

49 Bang, M. L. et al. Glial M6B stabilizes the axonal membrane at peripheral nodes of Ranvier. Glia 66, 801–812, doi:10.1002/glia.23285 (2018).

50 Banerjee, S. A. & Patterson, P. H. Schwann cell CD9 expression is regulated by axons. Mol Cell Neurosci 6, 462–473, doi:10.1006/mcne.1995.1034 (1995).

51 Schira, J. et al. Secretome analysis of nerve repair mediating Schwann cells reveals Smad-dependent trophism. Faseb j 33, 4703–4715, doi:10.1096/fj.201801799R (2019).

52 Ranjan, M. & Hudson, L. D. Regulation of tyrosine phosphorylation and protein tyrosine phosphatases during oligodendrocyte differentiation. Mol Cell Neurosci 7, 404–418, doi:10.1006/mcne.1996.0029 (1996).

53 Zhou, Z., Zhang, N., Shi, P. & Xie, J. Mechanism of miR-148b inhibiting cell proliferation and migration of Schwann cells by regulating CALR. Artif Cells Nanomed Biotechnol 47, 1978–1983, doi:10.1080/21691401.2019.1609008 (2019).

54 Zhang, Z. et al. Fibroblast-derived tenascin-C promotes Schwann cell migration through β1-integrin dependent pathway during peripheral nerve regeneration. Glia 64, 374–385, doi:10.1002/glia.22934 (2016).

55 Guo, S. K., Shen, M. F., Yao, H. W. & Liu, Y. S. Enhanced Expression of TGFBI Promotes the Proliferation and Migration of Glioma Cells. Cell Physiol Biochem 49, 1097–1109, doi:10.1159/000493293 (2018).

56 Naba, A. et al. The extracellular matrix: Tools and insights for the “omics” era. Matrix Biol 49, 10–24, doi:10.1016/j.matbio.2015.06.003 (2016).

57 Mazuelas, H., Carrió, M. & Serra, E. Modeling tumors of the peripheral nervous system associated with Neurofibromatosis type 1: Reprogramming plexiform neurofibroma cells. Stem Cell Res 49, 102068, doi:10.1016/j.scr.2020.102068 (2020).

58 Brosseau, J. P. et al. Human cutaneous neurofibroma matrisome revealed by single-cell RNA sequencing. Acta Neuropathol Commun 9, 11, doi:10.1186/s40478-020-01103-4 (2021).

59 Getting, S. J. et al. The link module from human TSG-6 inhibits neutrophil migration in a hyaluronan- and inter-alpha-inhibitor-independent manner. J Biol Chem 277, 51068–51076, doi:10.1074/jbc.M205121200 (2002).

60 Lin, Q. M. et al. Mesenchymal stem cells transplantation suppresses inflammatory responses in global cerebral ischemia: contribution of TNF-α-induced protein 6. Acta Pharmacol Sin 34, 784–792, doi:10.1038/aps.2012.199 (2013).

61 Nagyeri, G. et al. TSG-6 protein, a negative regulator of inflammatory arthritis, forms a ternary complex with murine mast cell tryptases and heparin. J Biol Chem 286, 23559–23569, doi:10.1074/jbc.M111.222026 (2011).

62 Wan, Y. M. et al. TSG-6 Inhibits Oxidative Stress and Induces M2 Polarization of Hepatic Macrophages in Mice With Alcoholic Hepatitis via Suppression of STAT3 Activation. Front Pharmacol 11, 10, doi:10.3389/fphar.2020.00010 (2020).

63 Mittal, M. et al. TNFα-stimulated gene-6 (TSG6) activates macrophage phenotype transition to prevent inflammatory lung injury. Proc Natl Acad Sci U S A 113, E8151–e8158, doi:10.1073/pnas.1614935113 (2016).

64 Chen, P. C. et al. Prostate cancer-derived CCN3 induces M2 macrophage infiltration and contributes to angiogenesis in prostate cancer microenvironment. Oncotarget 5, 1595–1608, doi:10.18632/oncotarget.1570 (2014).

65 Seki, E. et al. CCR2 promotes hepatic fibrosis in mice. Hepatology 50, 185–197, doi:10.1002/hep.22952 (2009).

66 Karlmark, K. R. et al. Hepatic recruitment of the inflammatory Gr1+ monocyte subset upon liver injury promotes hepatic fibrosis. Hepatology 50, 261–274, doi:10.1002/hep.22950 (2009).

67 Robinson, S. C., Scott, K. A. & Balkwill, F. R. Chemokine stimulation of monocyte matrix metalloproteinase-9 requires endogenous TNF-alpha. Eur J Immunol 32, 404–412, doi:10.1002/1521-4141(200202)32:2<404::Aid-immu404>3.0.Co;2-x (2002).

68 Stratton, J. A. et al. Macrophages Regulate Schwann Cell Maturation after Nerve Injury. Cell Rep 24, 2561-2572.e2566, doi:10.1016/j.celrep.2018.08.004 (2018).

69 Yasuoka, H., Jukic, D. M., Zhou, Z., Choi, A. M. & Feghali-Bostwick, C. A. Insulin-like growth factor binding protein 5 induces skin fibrosis: A novel murine model for dermal fibrosis. Arthritis Rheum 54, 3001–3010, doi:10.1002/art.22084 (2006).

70 Yasuoka, H. et al. Insulin-like growth factor-binding protein-5 induces pulmonary fibrosis and triggers mononuclear cellular infiltration. Am J Pathol 169, 1633–1642, doi:10.2353/ajpath.2006.060501 (2006).

71 Yasuoka, H., Yamaguchi, Y. & Feghali-Bostwick, C. A. The pro-fibrotic factor IGFBP-5 induces lung fibroblast and mononuclear cell migration. Am J Respir Cell Mol Biol 41, 179–188, doi:10.1165/rcmb.2008-0211OC (2009).

72 Min, H. K. et al. Suppression of IGF binding protein-3 by palmitate promotes hepatic inflammatory responses. Faseb j 30, 4071–4082, doi:10.1096/fj.201600427R (2016).

73 Reinisch, C. M. et al. Rarefaction of the peripheral nerve network in diabetic patients is associated with a pronounced reduction of terminal Schwann cells. Diabetes Care 31, 1219–1221, doi:10.2337/dc07-1832 (2008).

74 Choi, K. et al. An inflammatory gene signature distinguishes neurofibroma Schwann cells and macrophages from cells in the normal peripheral nervous system. Sci Rep 7, 43315, doi:10.1038/srep43315 (2017).

75 Carroll, S. L. & Ratner, N. How does the Schwann cell lineage form tumors in NF1? Glia 56, 1590–1605, doi:10.1002/glia.20776 (2008).

76 Haider, N. et al. Transition of Macrophages to Fibroblast-Like Cells in Healing Myocardial Infarction. J Am Coll Cardiol 74, 3124–3135, doi:10.1016/j.jacc.2019.10.036 (2019).

77 Thiery, J. P., Acloque, H., Huang, R. Y. & Nieto, M. A. Epithelial-mesenchymal transitions in development and disease. Cell 139, 871–890, doi:10.1016/j.cell.2009.11.007 (2009).

78 Kim, Y. et al. The MMP-9/TIMP-1 axis controls the status of differentiation and function of myelin-forming Schwann cells in nerve regeneration. PLoS One 7, e33664, doi:10.1371/journal.pone.0033664 (2012).

79 Hao, Y. et al. Integrated analysis of multimodal single-cell data. Cell, doi:10.1016/j.cell.2021.04.048 (2021).

80 Ahlmann-Eltze, C. & Huber, W. glmGamPoi: fitting Gamma-Poisson generalized linear models on single cell count data. Bioinformatics 36, 5701–5702, doi:10.1093/bioinformatics/btaa1009 (2021).

81 Hafemeister, C. & Satija, R. Normalization and variance stabilization of single-cell RNA-seq data using regularized negative binomial regression. Genome Biology 20, 296, doi:10.1186/s13059-019-1874-1 (2019).

82 Trapnell, C. et al. The dynamics and regulators of cell fate decisions are revealed by pseudotemporal ordering of single cells. Nat Biotechnol 32, 381–386, doi:10.1038/nbt.2859 (2014).

83 Qiu, X. et al. Reversed graph embedding resolves complex single-cell trajectories. Nat Methods 14, 979–982, doi:10.1038/nmeth.4402 (2017).

84 Cao, J. et al. The single-cell transcriptional landscape of mammalian organogenesis. Nature 566, 496–502, doi:10.1038/s41586-019-0969-x (2019).

85 Traag, V. A., Waltman, L. & van Eck, N.J. From Louvain to Leiden: guaranteeing well-connected communities. Sci Rep 9, 5233, doi:10.1038/s41598-019-41695-z (2019).

86 Levine, J. H. et al. Data-Driven Phenotypic Dissection of AML Reveals Progenitor-like Cells that Correlate with Prognosis. Cell 162, 184–197, doi:10.1016/j.cell.2015.05.047 (2015).

87 Zhou, Y. et al. Metascape provides a biologist-oriented resource for the analysis of systems-level datasets. Nat Commun 10, 1523, doi:10.1038/s41467-019-09234-6 (2019).

## References

1 Tabib, T., Morse, C., Wang, T., Chen, W. & Lafyatis, R. SFRP2/DPP4 and FMO1/LSP1 Define Major Fibroblast Populations in Human Skin. J Invest Dermatol 138, 802–810, doi:10.1016/j.jid.2017.09.045 (2018).

## References

1 Muhl, L. et al. Single-cell analysis uncovers fibroblast heterogeneity and criteria for fibroblast and mural cell identification and discrimination. Nat Commun 11, 3953, doi:10.1038/s41467-020-17740-1 (2020).

2 Vorstandlechner, V. et al. Deciphering the functional heterogeneity of skin fibroblasts using single-cell RNA sequencing. Faseb j 34, 3677–3692, doi:10.1096/fj.201902001RR (2020).

3 Tabib, T., Morse, C., Wang, T., Chen, W. & Lafyatis, R. SFRP2/DPP4 and FMO1/LSP1 Define Major Fibroblast Populations in Human Skin. J Invest Dermatol 138, 802–810, doi:10.1016/j.jid.2017.09.045 (2018).

4 Wolbert, J. et al. Redefining the heterogeneity of peripheral nerve cells in health and autoimmunity. Proc Natl Acad Sci U S A 117, 9466–9476, doi:10.1073/pnas.1912139117 (2020).

5 Voigt, A. P. et al. Single-cell transcriptomics of the human retinal pigment epithelium and choroid in health and macular degeneration. Proc Natl Acad Sci U S A 116, 24100–24107, doi:10.1073/pnas.1914143116 (2019).

6 Finnegan, A. et al. Single-Cell Transcriptomics Reveals Spatial and Temporal Turnover of Keratinocyte Differentiation Regulators. Front Genet 10, 775, doi:10.3389/fgene.2019.00775 (2019).

7 Saichi, M. et al. Single-cell RNA sequencing of blood antigen-presenting cells in severe COVID-19 reveals multi-process defects in antiviral immunity. Nat Cell Biol 23, 538–551, doi:10.1038/s41556-021-00681-2 (2021).

8 Elsner, R. A., Ernst, D. N. & Baumgarth, N. Single and coexpression of CXCR4 and CXCR5 identifies CD4 T helper cells in distinct lymph node niches during influenza virus infection. J Virol 86, 7146–7157, doi:10.1128/jvi.06904-x11 (2012).

9 Villani, A. C. et al. Single-cell RNA-seq reveals new types of human blood dendritic cells, monocytes, and progenitors. Science 356, doi:10.1126/science.aah4573 (2017).

