## Supplementary Figures and Tables for "Schwann cells contribute to keloid formation"

### Supplementary Figure 1

(a)

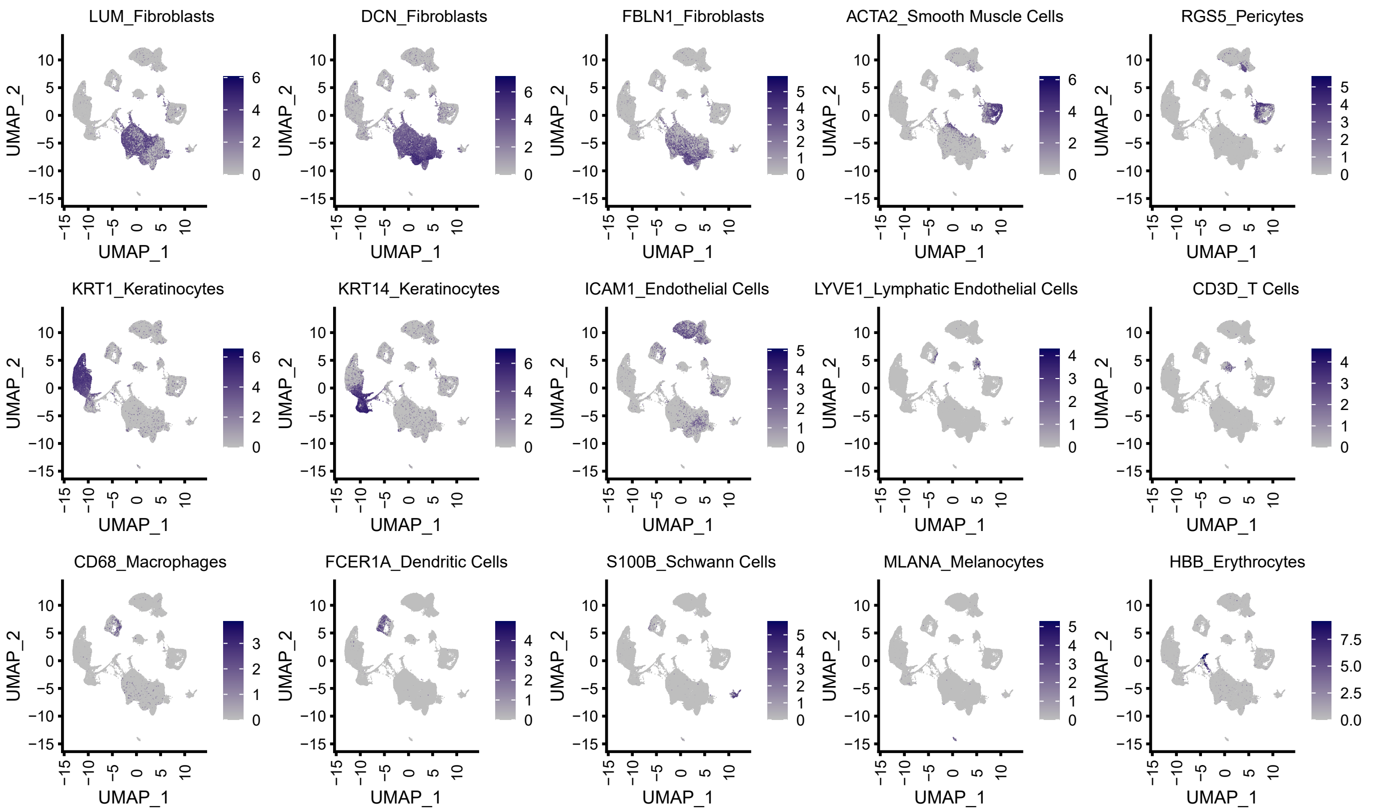


(b)

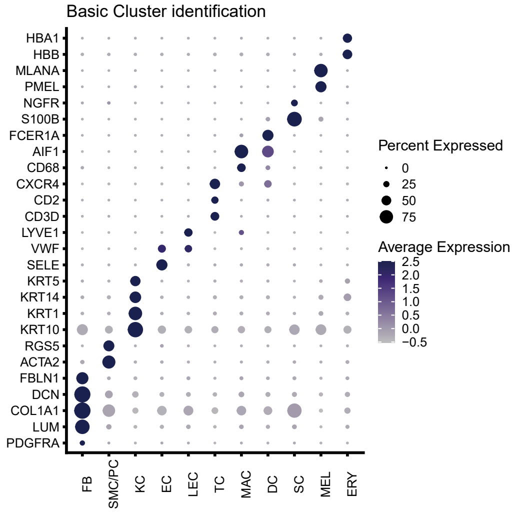

Supplementary Figure 2

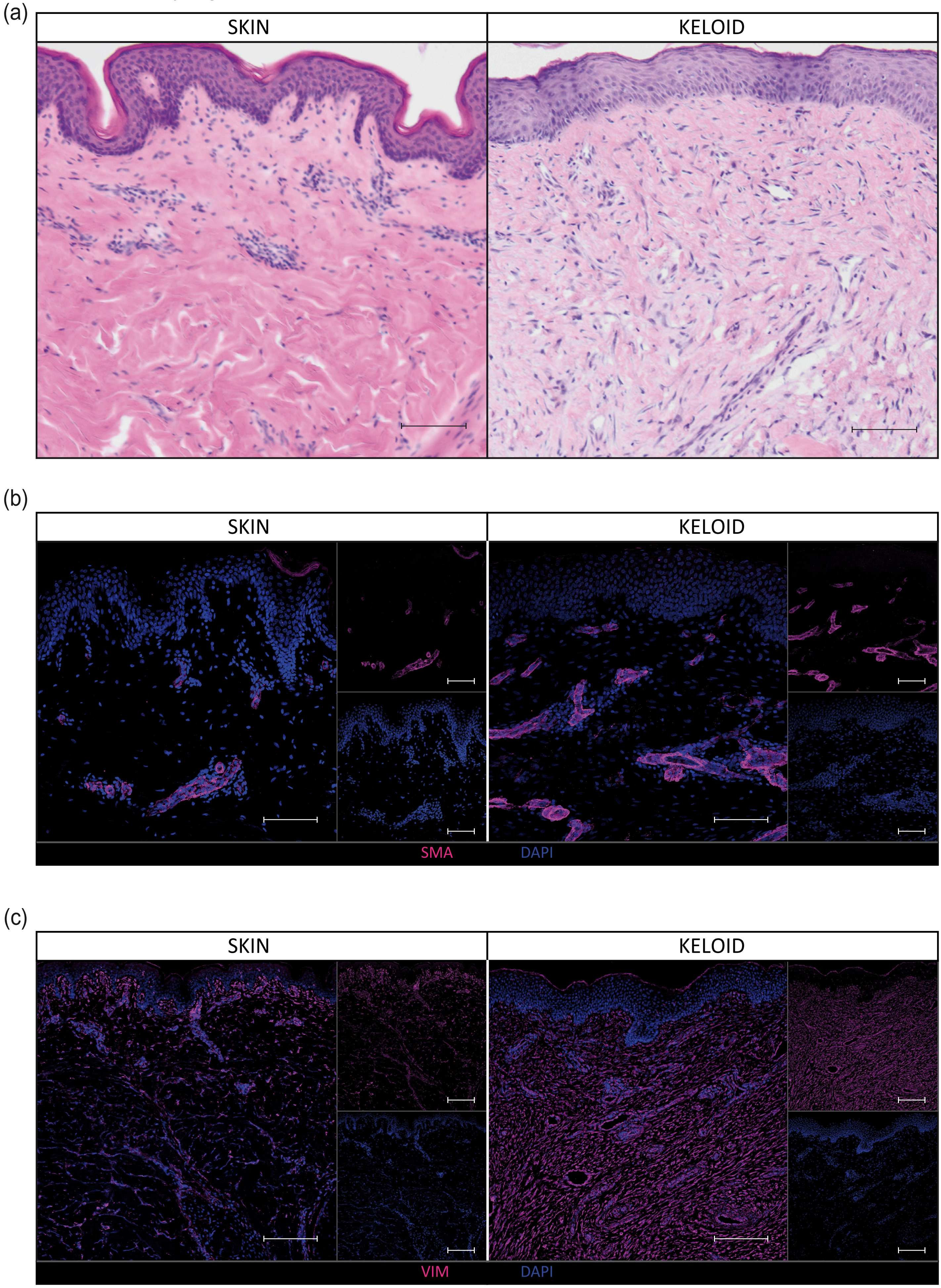

Supplementary Figure 3

(a)

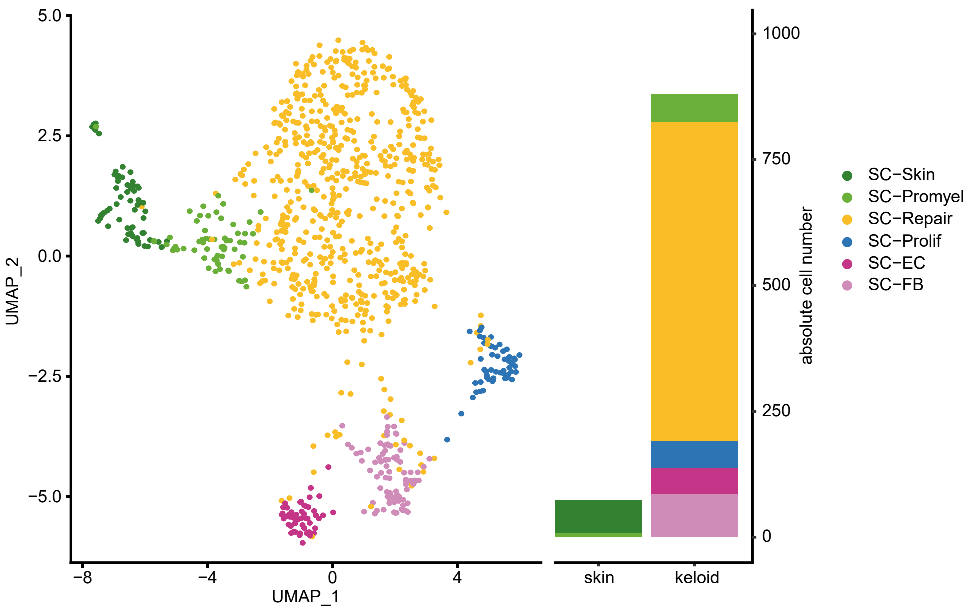

Supplementary Figure 4

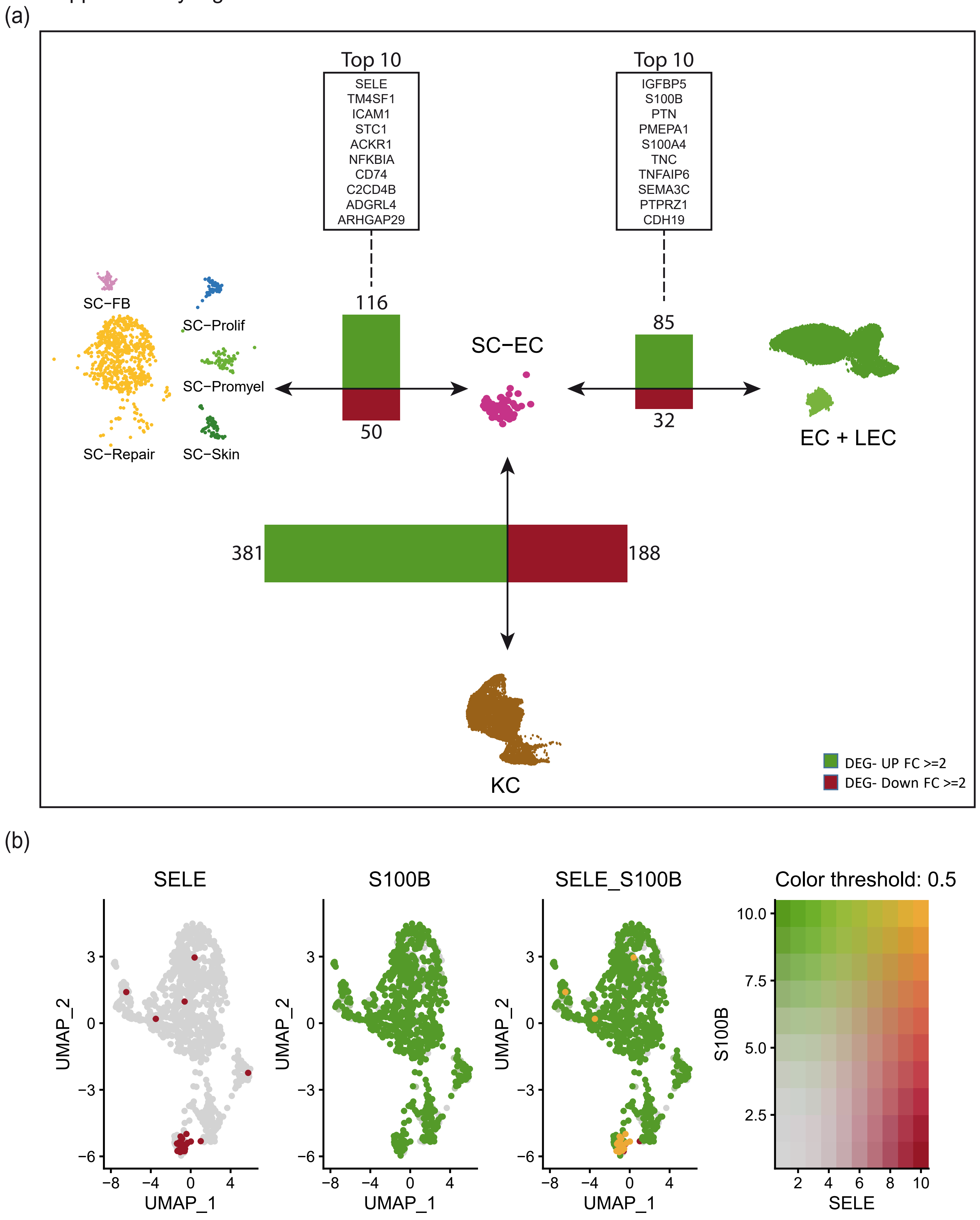

Supplementary Figure 5

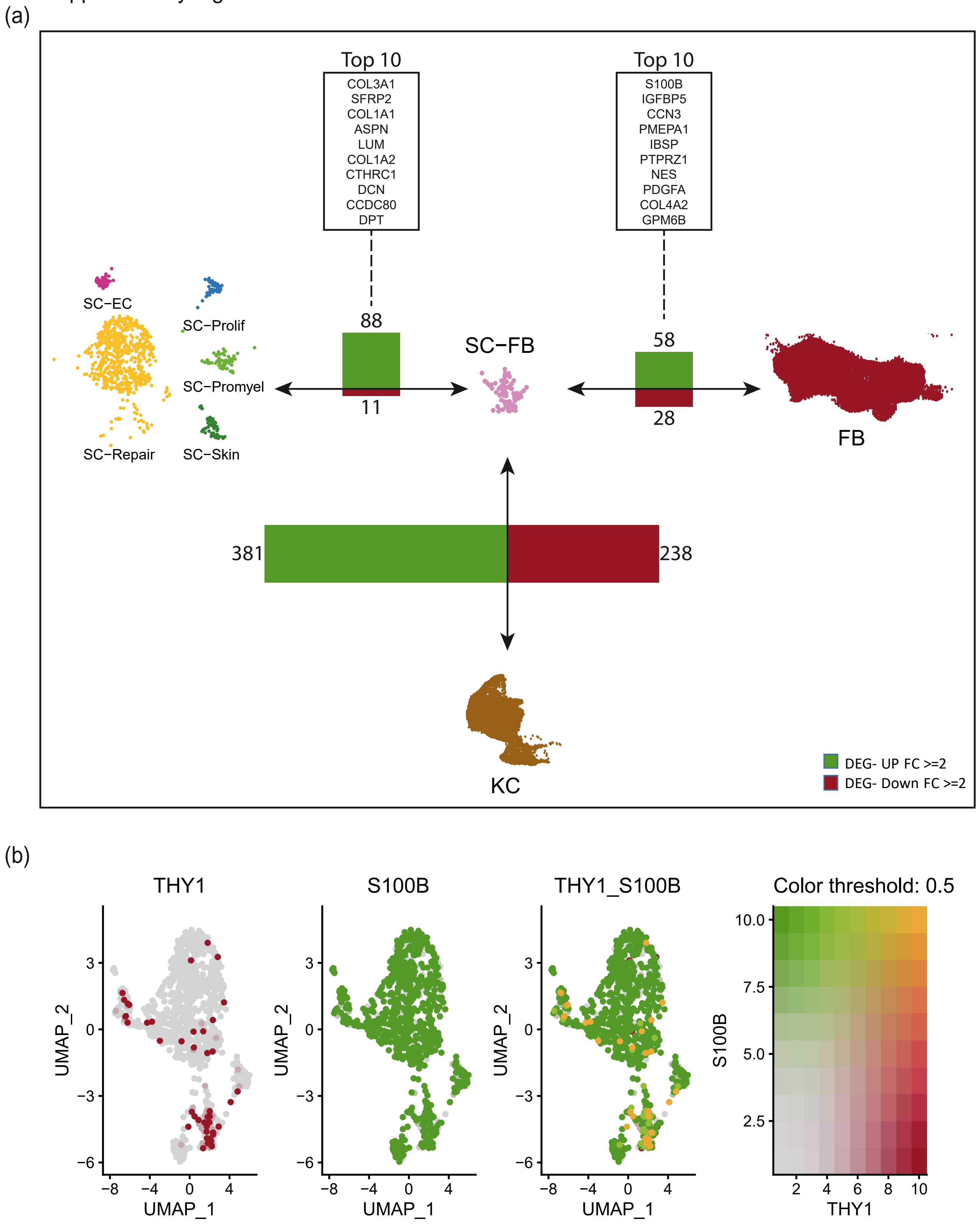

### Supplementary Figure 6

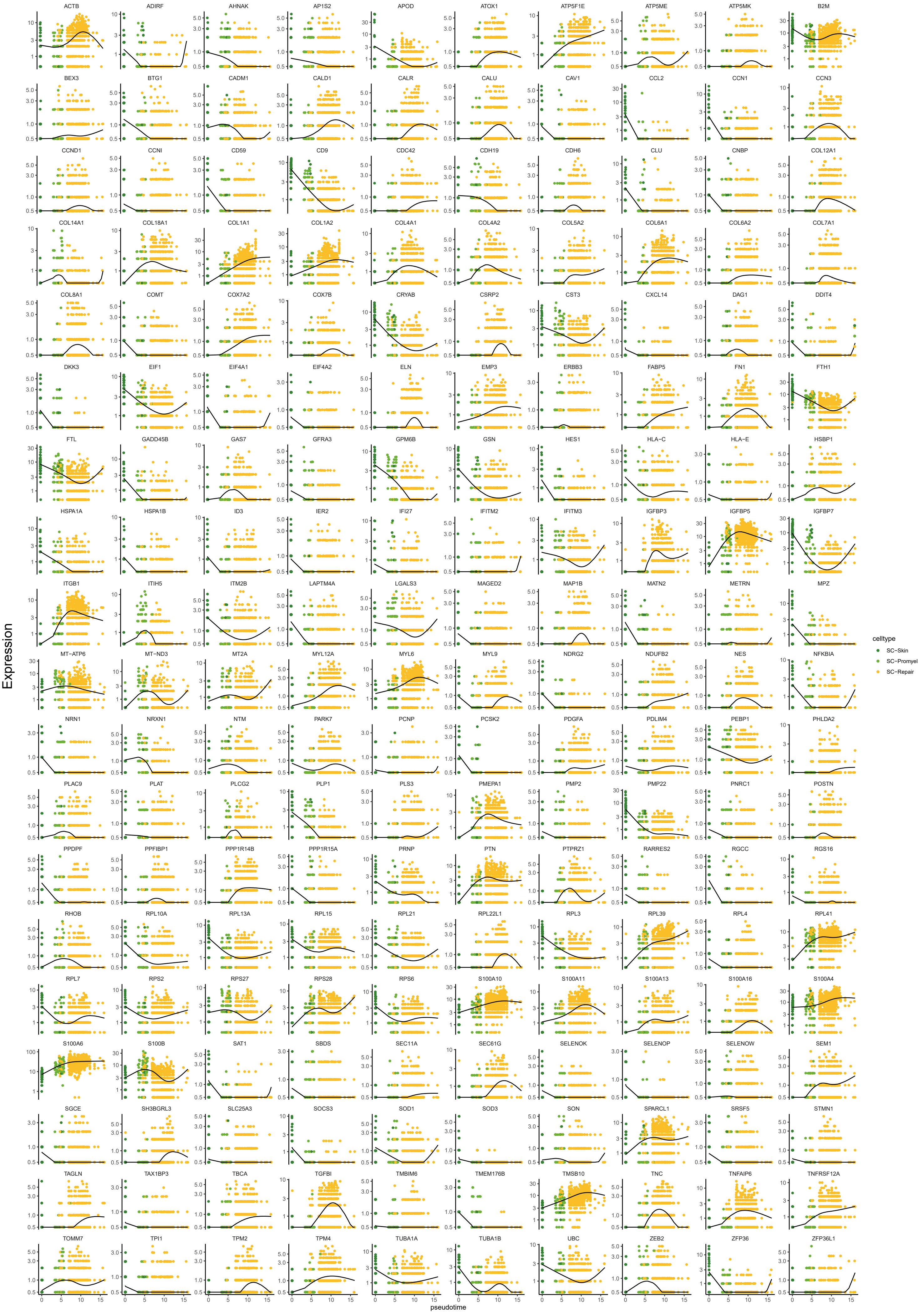

### Supplementary Figure 7

(a)

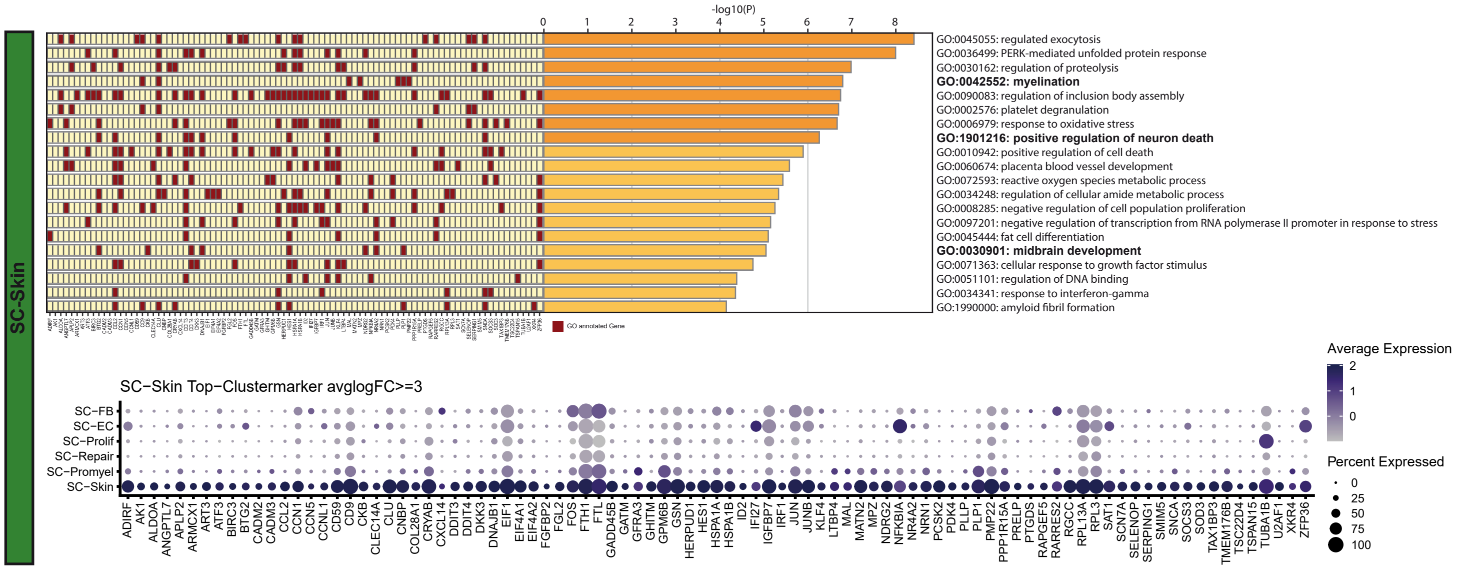


(b)

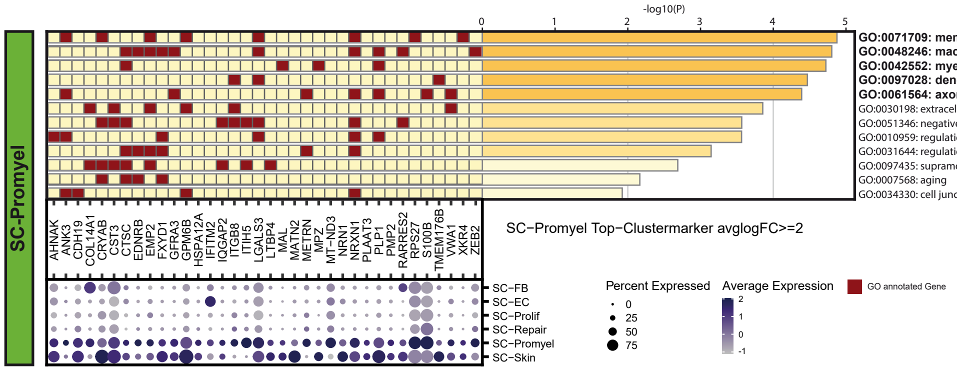

### Supplementary Figure 8

(a)

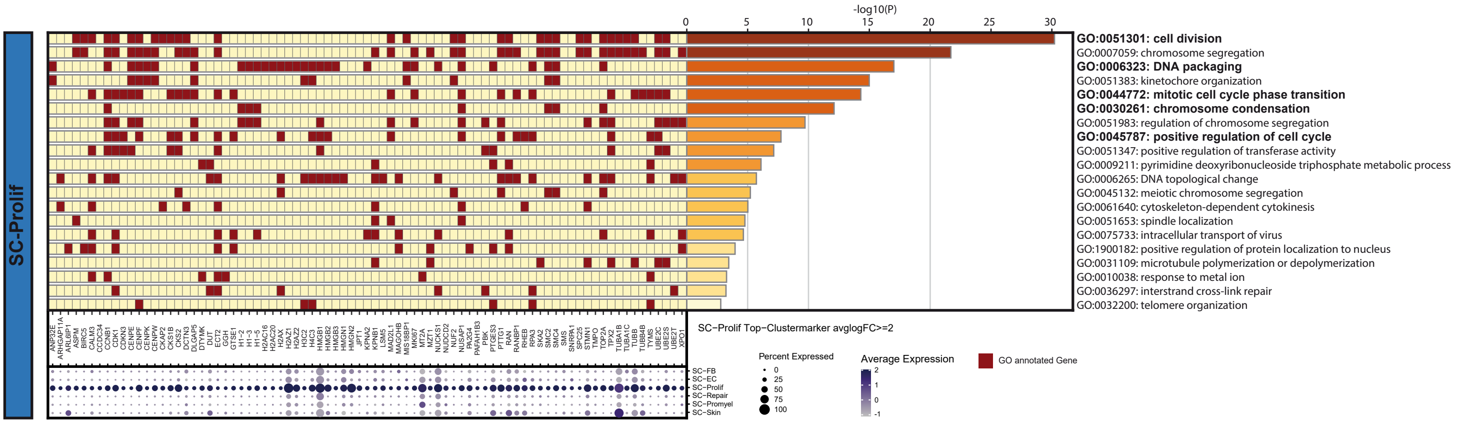


(b)

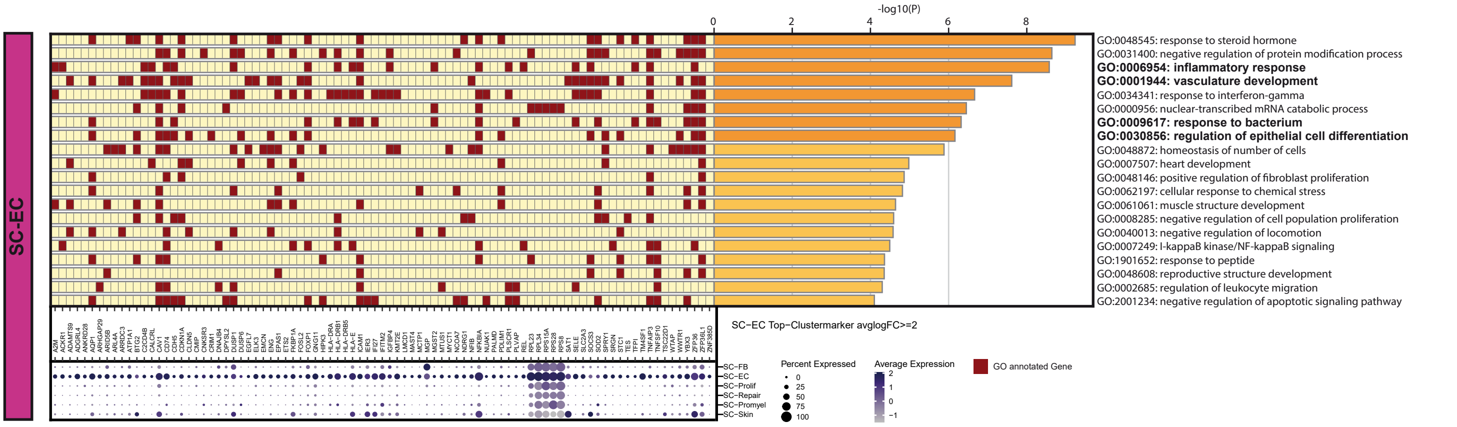


(c)

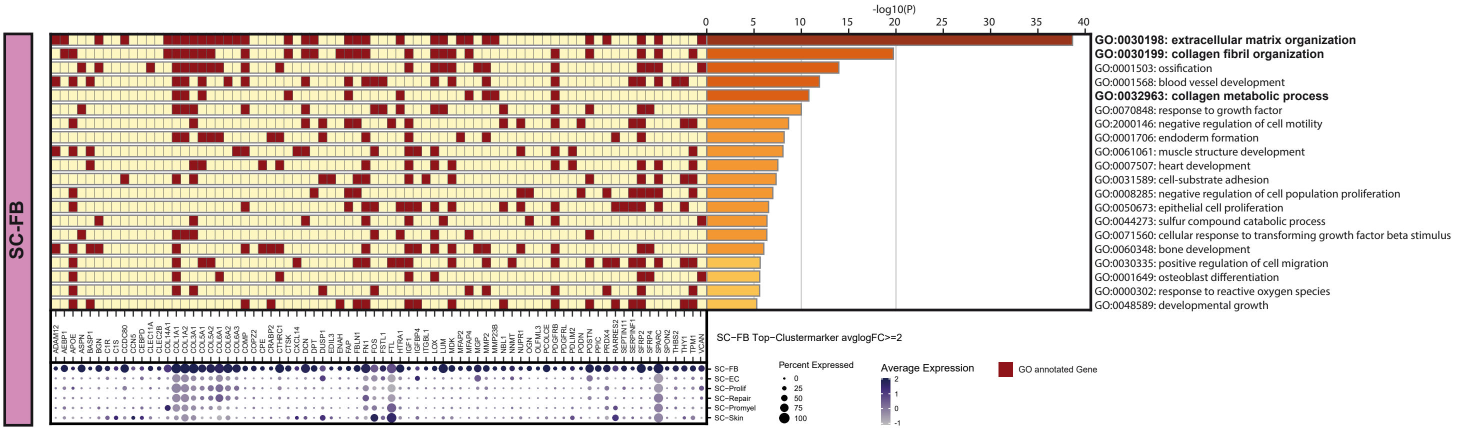

### Supplementary Figure 9

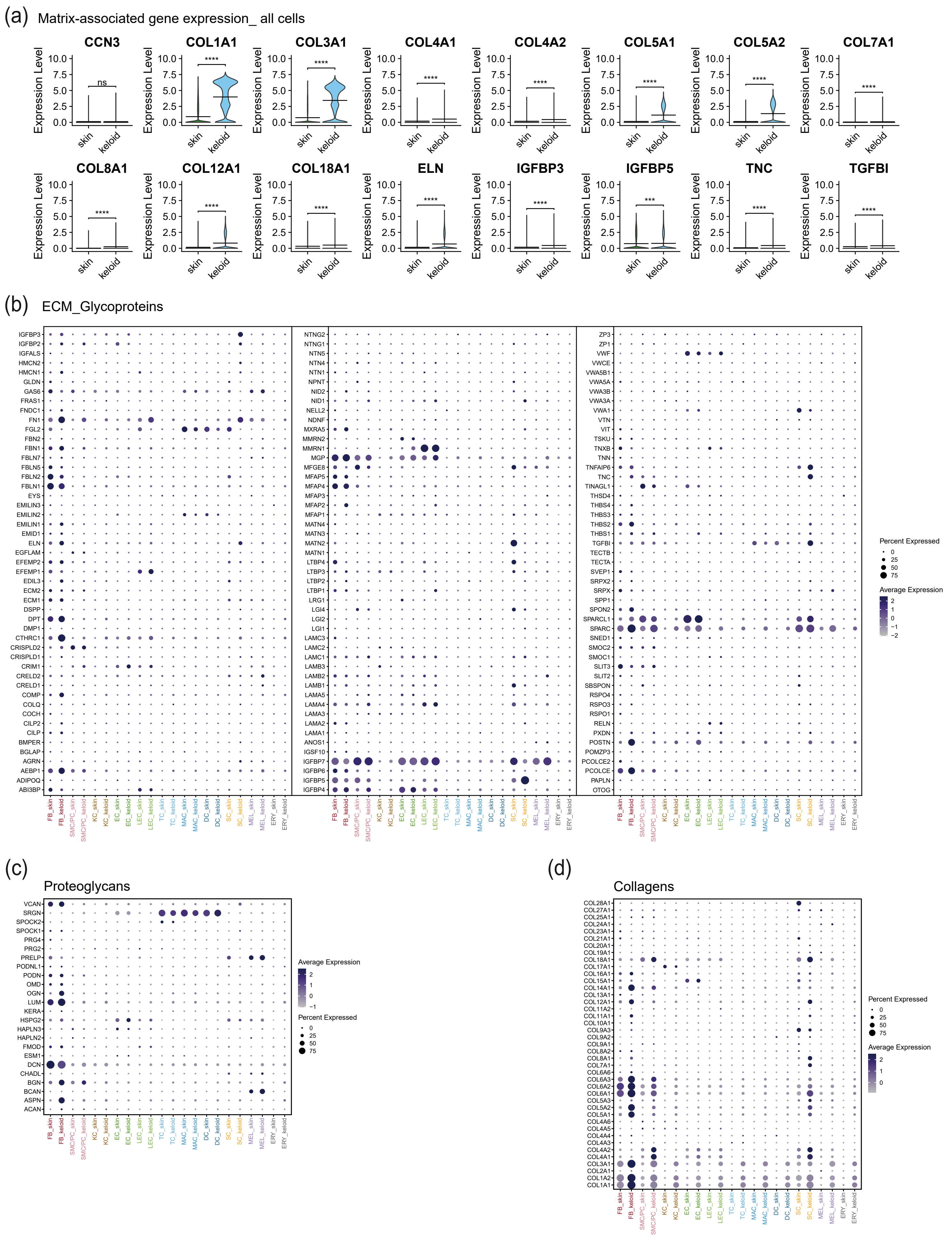

Supplementary Figure 10

(a) ECM_Regulators

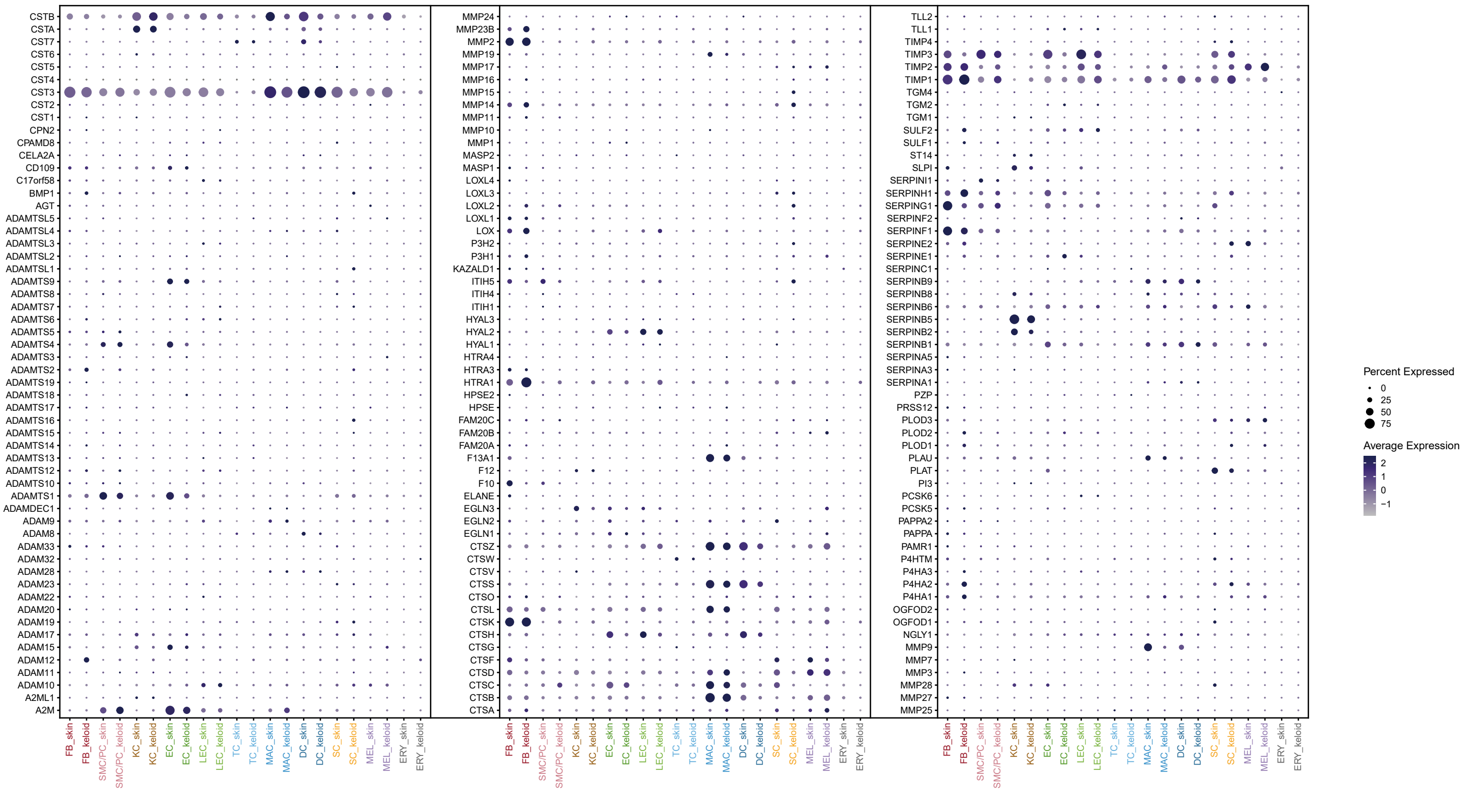


(b) ECM_affiliated_Proteins

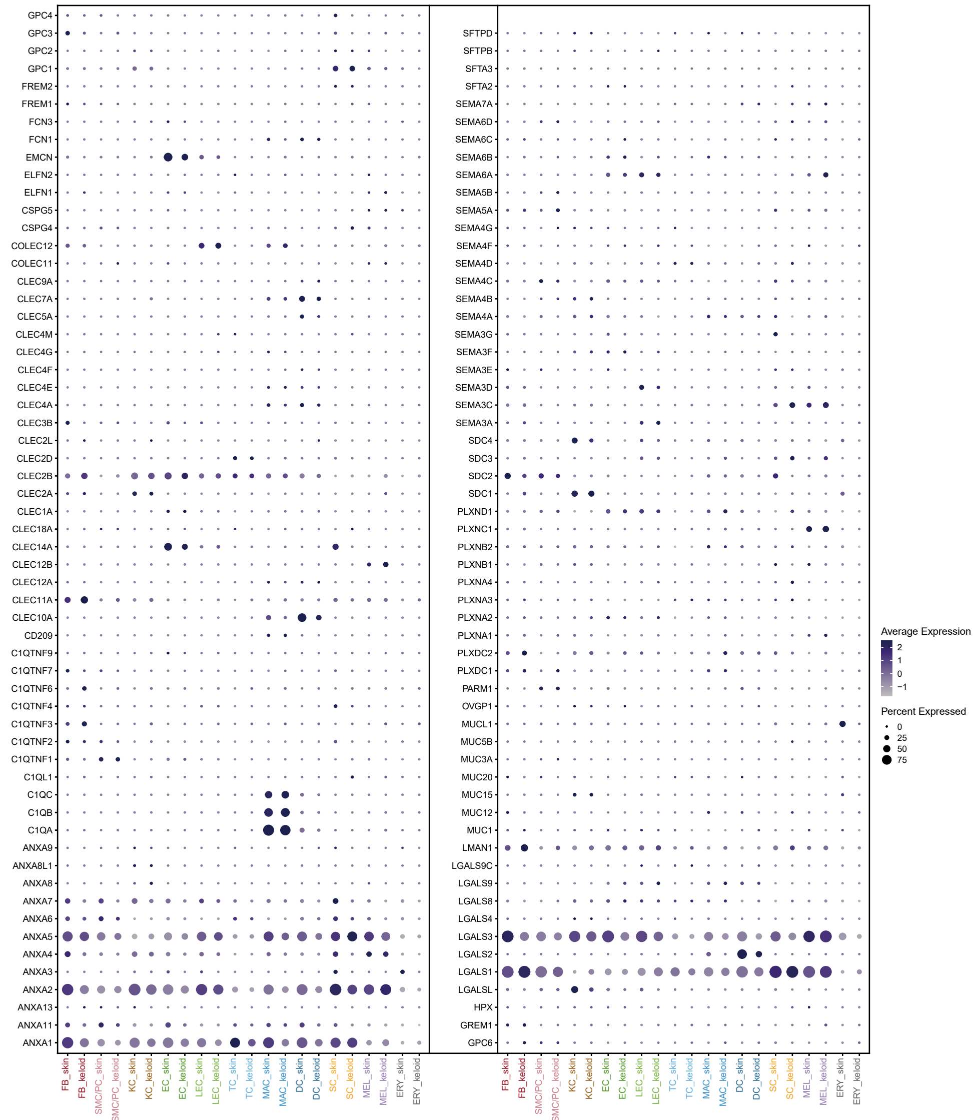

### Supplementary Figure 11

(a)

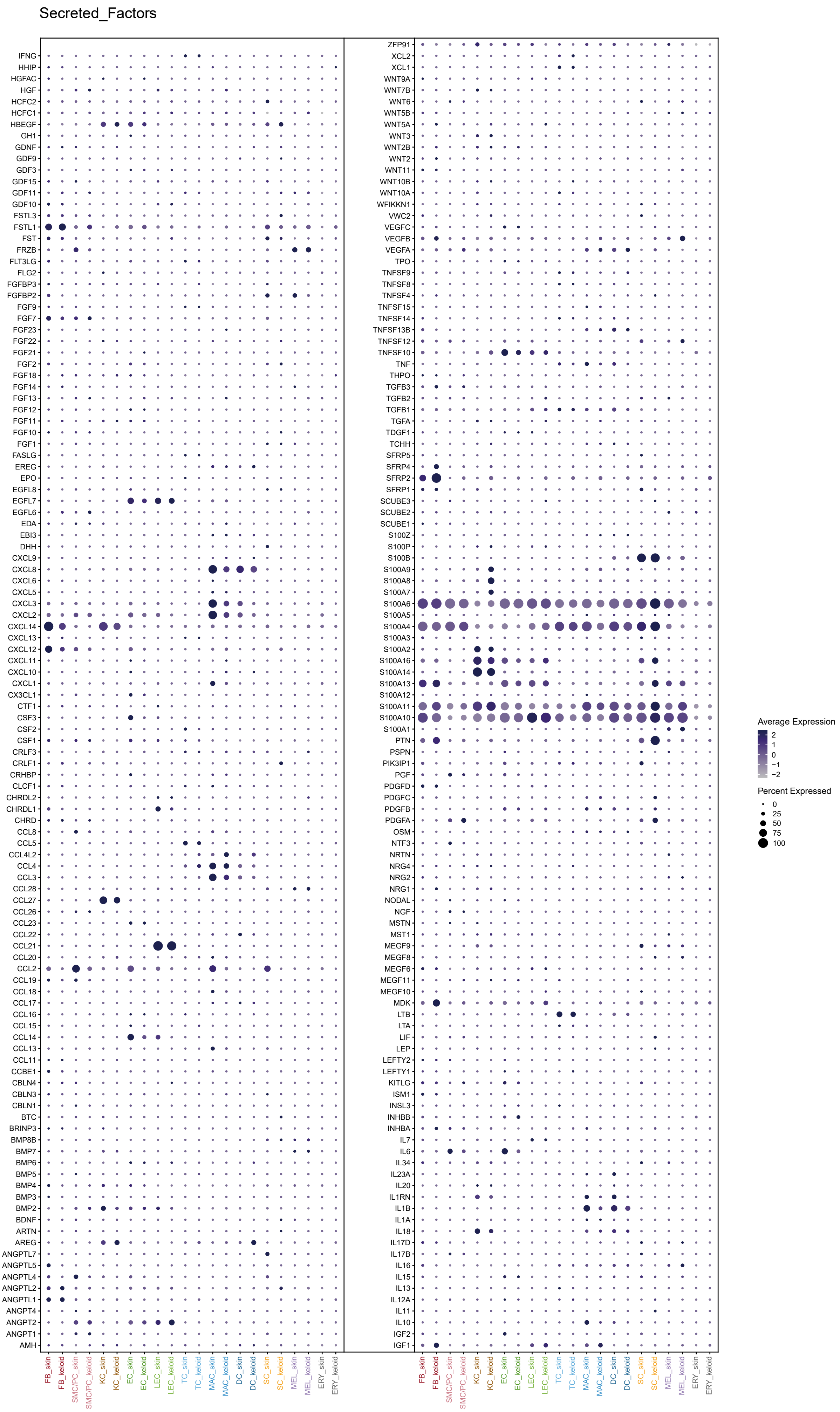

Supplementary Figure 12

(a) Inflammation-associated gene expression_ all cells

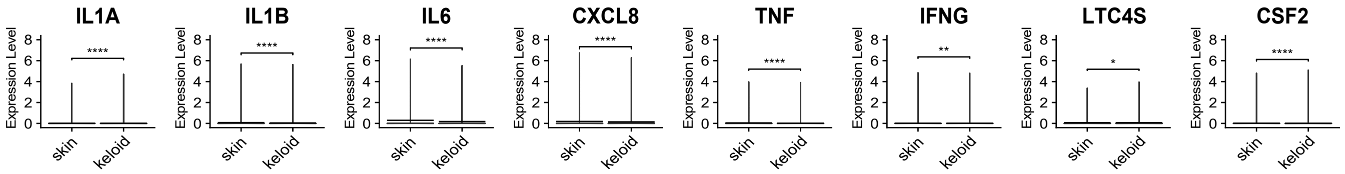


(b) Immunology-associated genes in cells of skin and keloid

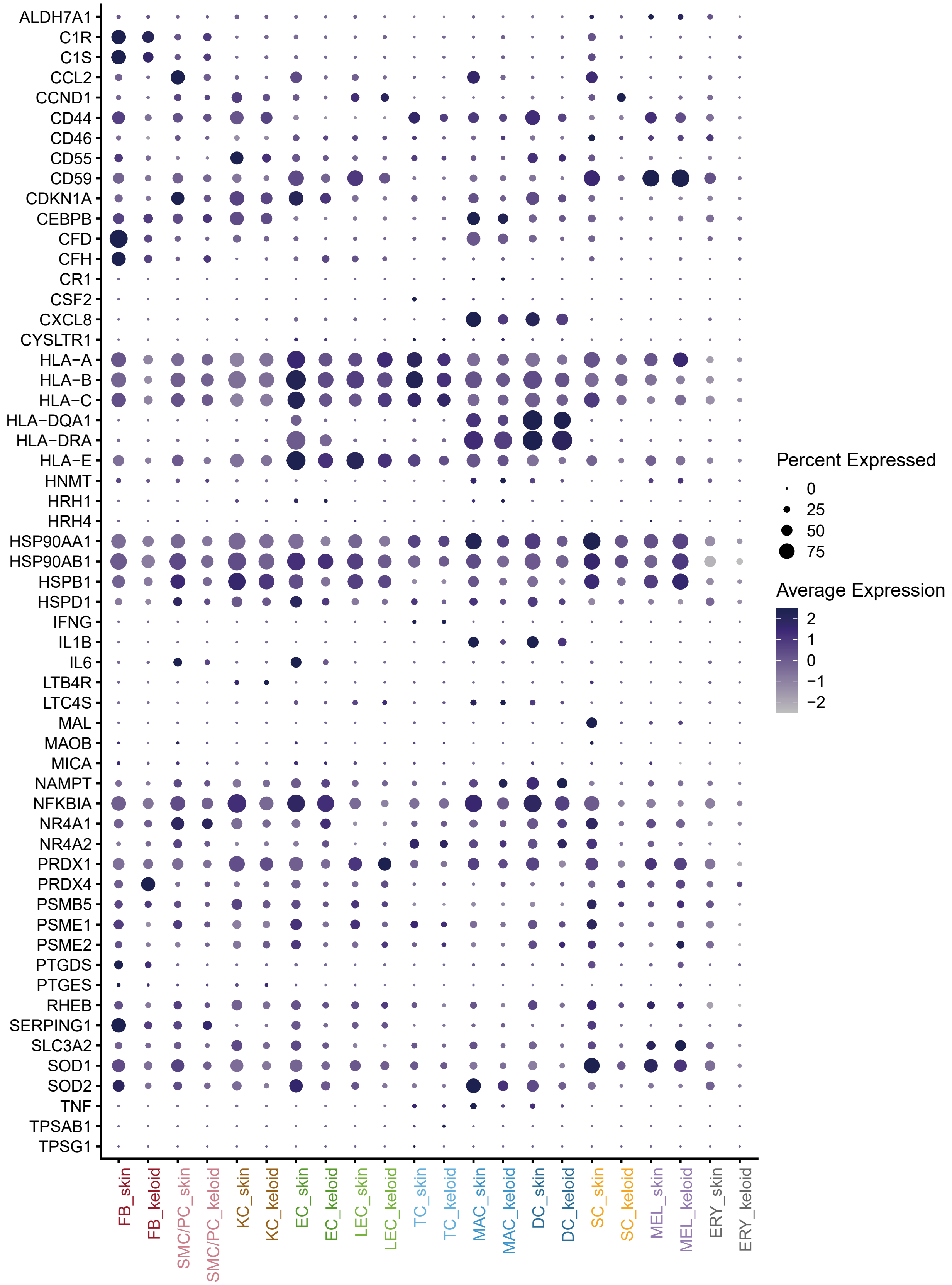

Supplementary Figure 13

(a)

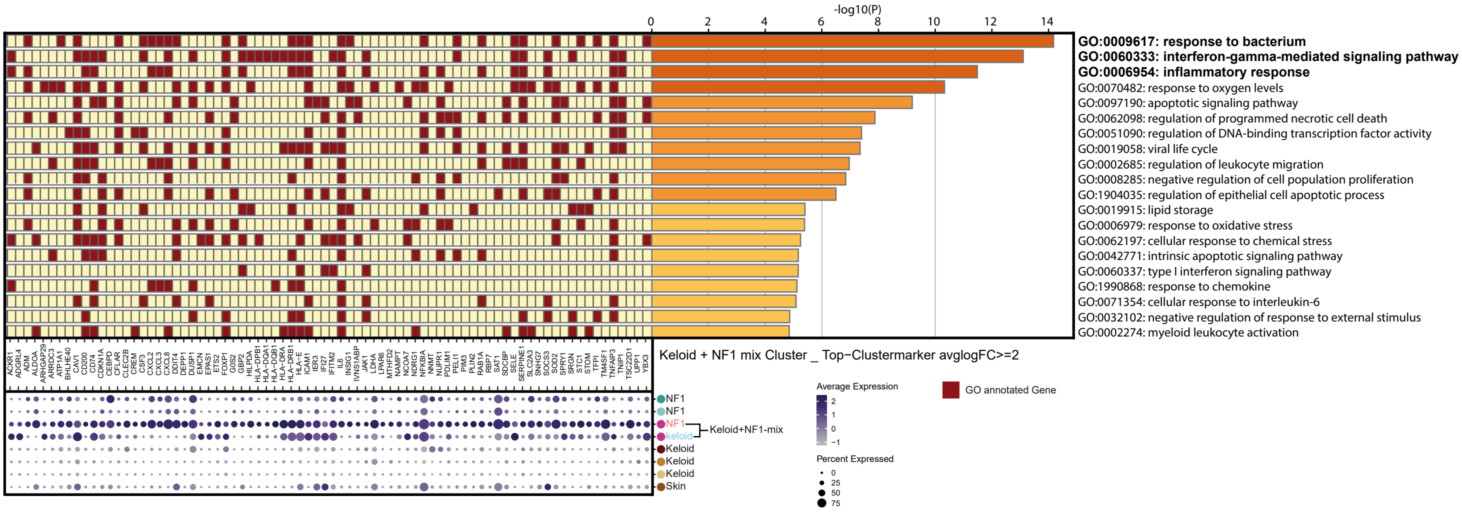

Supplementary Figure 14

(a)

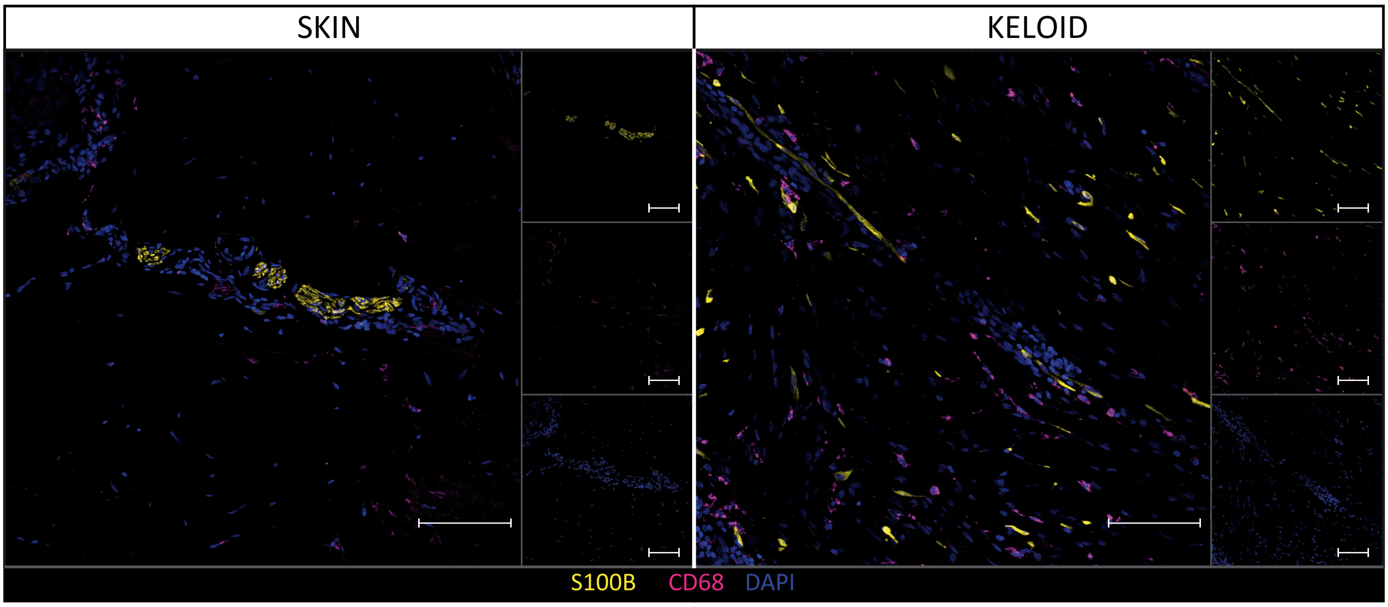


(b)

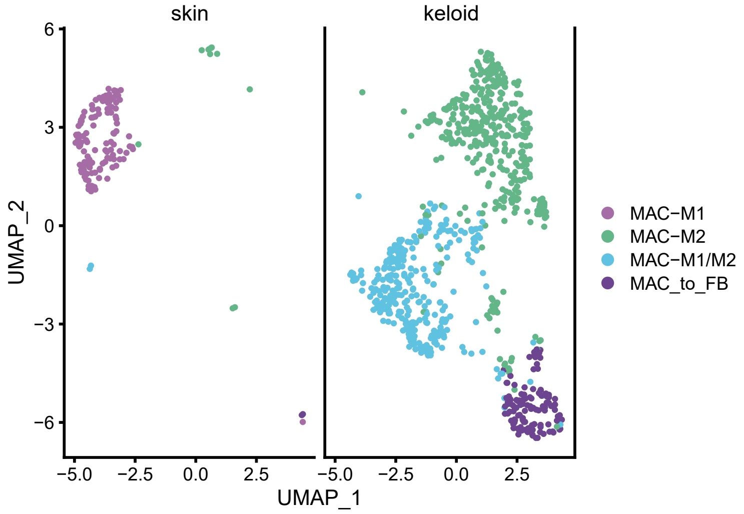

Supplementary Figure 15

(a)

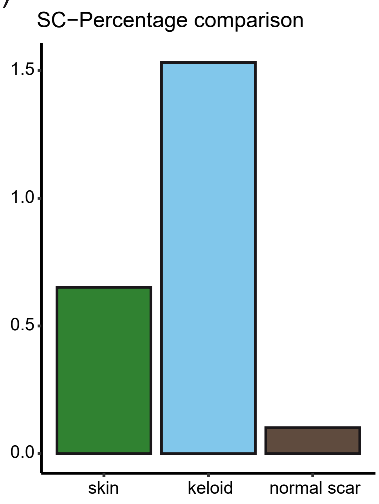

Supplementary Figure 16

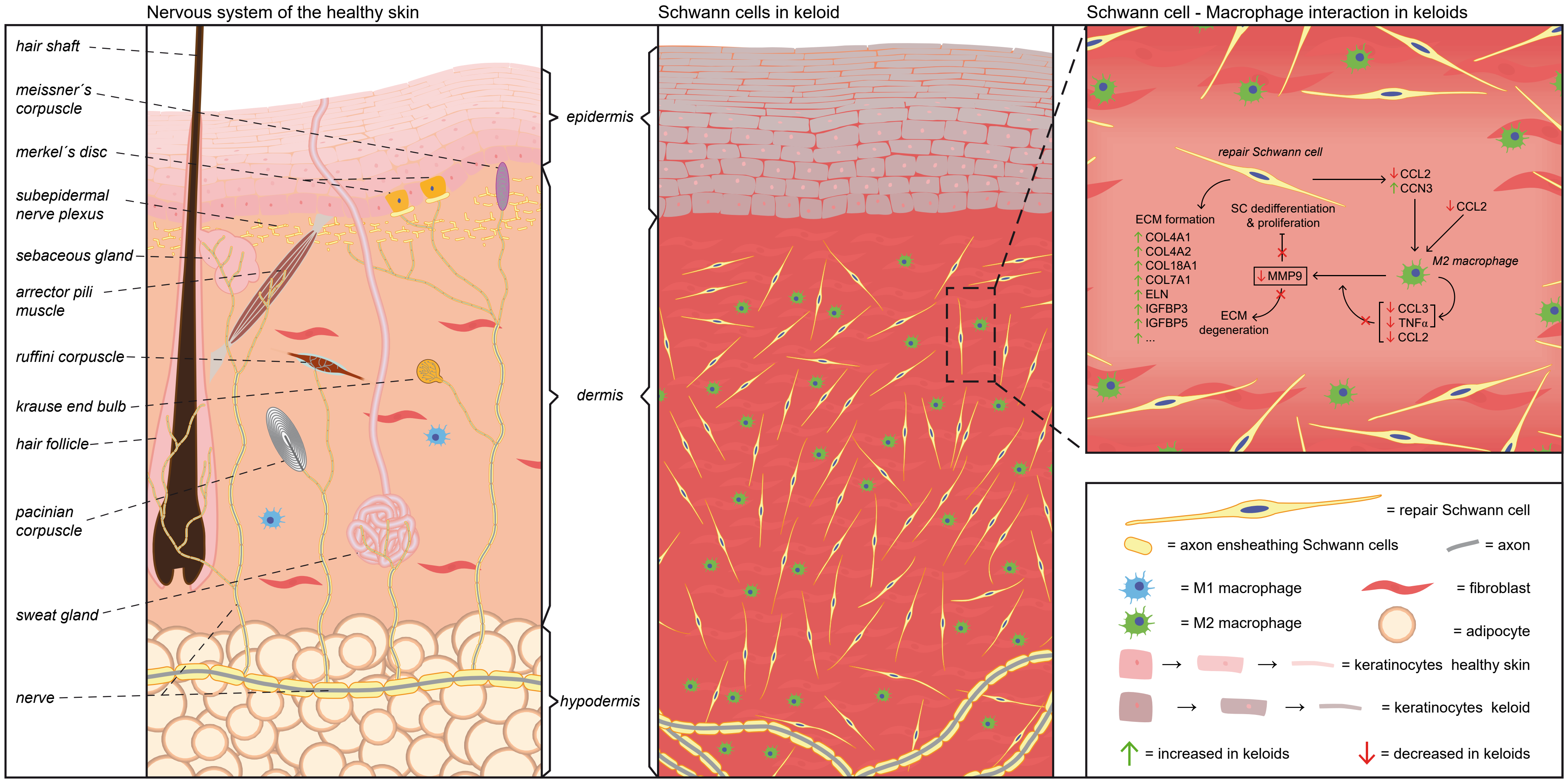

Supplementary Table 1 – donor information

| Type | ID | Site | Sex | Race | Source |
|---|---|---|---|---|---|
| normal skin | skin_1 | Forearm | Male | Caucasian | Tabib[1] |
| normal skin | skin_2 | Forearm | Male | Caucasian | Tabib[1] |
| normal skin | skin_3 | Forearm | Female | Caucasian | Tabib[1] |
| normal skin | skin_4 | Forearm | Female | Asian | Tabib[1] |
| normal skin | skin_5 | Forearm | Female | Caucasian | Tabib[1] |
| normal skin | skin_6 | Forearm | Male | Caucasian | Tabib[1] |
| normal skin | skin_7 | Abdomen | Male | Caucasian | Direder |
| Keloid | keloid_1 | Sternum | Female | Caucasian | Direder |
| Keloid | keloid_2 | Earlobe | Female | African | Direder |
| Keloid | keloid_3L | Earlobe - left | Male | Caucasian | Direder |
| Keloid | keloid_3R | Earlobe - right | Male | Caucasian | Direder |
| normal scar | scar1 | Abdomen | Female | Caucasian | Direder |
| normal scar | scar2 | Abdomen | Female | Caucasian | Direder |
| normal scar | scar3 | Abdomen | Male | Caucasian | Direder |

#### Supplementary Table 2 - marker genes information

| celltype | abbreviation | marker gene |
|---|---|---|
| Fibroblast | FB | PDGFRA[1], LUM[2], COL1A1[3,4], DCN[1], FBLN1[5] |
| Smooth muscle cell or pericyte | SMC/PC | ACTA2[2,4], RGS5[2,4,5] |
| Keratinocyte | KC | KRT10[6], KRT1[2,6], KRT14[2,6], KRT5[6] |
| Endothelial cell | EC | SELE[2], VWF[5] |
| Lymphatic Endothelial cell | LEC | VWF[5], LYVE1[2,4] |
| T-cell | TC | CD3D[4,7], CD2[5], CXCR4[8] |
| Macrophage | MAC | CD68[4], AIF1[2] |
| Dendritic cell | DC | AIF1[2], FCER1A[7,9] |
| Schwann cell | SC | S100B[4], NGFR[4,5] |
| Melanocyte | MEL | PMEL[2,5], MLANA[5] |
| Erythrocyte | ERY | HBB[7], HBA1[7] |

### Supplementary Table 3 – antibody information

| 1st Antibodies | | | | | |
|---|---|---|---|---|---|
| Antigen | Species | catalog No | company | dilution | comment |
| S100 | rabbit | #Z0311 | DAKO | ready to use | 4 hr, RT |
| NGFR | mouse | #sc-13577 | SantaCruz | 1:100 | o.n., 4°C |
| NGFR | rabbit | #8238 | CellSignaling | 1:300 | 4 hr, RT |
| SOX10 | mouse | #sc-365692 | SantaCruz | 1:33 | o.n., 4°C |
| Nestin | mouse | #MAB5326 | Millipore | 1:200 | o.n., 4°C |
| Ki67 | rabbit | #NB500-170 | Novus | 1:50 | o.n., 4°C |
| MBP | mouse | #ab62631 | Abcam | 1:300 | o.n., 4°C |
| CD31 | mouse | #M0823 | DAKO | 1:20 | 4 hr, RT |
| IGFBP-5 | goat | #AF875-SP | R&D | 1:50 | o.n., 4°C |
| JUN | rabbit | #9165S | CellSignaling | 1:300 | 4 hr, RT |
| CD90 | mouse | # 550402 | BD Pharmingen | 1:50 | 4 hr, RT |
| PGP9.5 | mouse | #7863-1004 | BioRad | 1:250 | o.n., 4°C |
| vimentin | chicken | #AB5733 | Merck Millipore | 1:300 | 4 hr, RT |
| SMA | rabbit | #14395-I-AP | Proteintech | 1:200 | 4 hr, RT |
| CD68 | mouse | #333809 | BioLegend | 1:50 | o.n., 4°C |
| 2nd Antibodies | | | | | |
| Antigen | Species | catalog No | company | dilution | comment |
| α ch DL650 | goat | #SA5-10073 | Invitrogen | 1:400 | 1 hr, RT |
| α rb AF488 | goat | #A32731 | Invitrogen | 1:600 | 1 hr, RT |
| α ch AF488 | goat | #A-11039 | Invitrogen | 1:400 | 1 hr, RT |
| α rb AF4594 | goat | #A11012 | Invitrogen | 1:400 | 1 hr, RT |
| α ms AF594 | donkey | #A21203 | Invitrogen | 1:400 | 1 hr, RT |
| α g AF546 | donkey | #A11056 | Invitrogen | 1:400 | 1 hr, RT |
